# An integrated analysis of amniotic fluid reveals a dynamic prenatal microbial landscape and regulatory axis

**DOI:** 10.1101/2025.07.29.25331497

**Authors:** M González-Rovira, JA Sainz-Bueno, L García-Díaz, C Martínez-Pancorbo, J Sánchez, G Gutiérrez, K Magoutas, A Mesías-Pérez, E Mellado, M Payne, C Sousa, ML Moreno

## Abstract

Challenging the sterile womb dogma, our study investigated amniotic fluid (AF) microbial colonization and its potential role in foetal immune development. We analysed microbial presence in 154 AF samples without clinical and analytical suspicion of AF infection using optimised bacterial culture methods and full-length 16S rRNA gene sequencing. Shotgun metagenomic sequencing was also performed on a subset of culture-positive samples, and antimicrobial peptide levels were also measured. Remarkably, 29.5% of amniocentesis-derived and 55.0% of caesarean-derived samples yielded positive cultures, predominantly *Bacillus*, *Cutibacterium*, *Micrococcus*, and *Staphylococcus* genera, with *Cutibacterium acnes* and *Staphylococcus epidermidis* being most common species. Both sequencing methods revealed a low-biomass, low-diversity microbial community with high inter-individual variability. Higher HBD-1 levels correlated with an absence of culturable bacteria or detectable microbial DNA. While sequencing showed no significant microbial differences based on gestational age or pregnancy outcome, bacterial species identified through culture varied with reproductive treatments. Of most interest, diamniotic twins showed intra-individual microbial discordance between sacs, directly challenging intrauterine homogeneity. Our findings indicate that viable bacteria and/or their DNA can transiently access the prenatal environment. This highlights the importance of a possible microbial balance in the prenatal setting, suggesting a novel perspective of a potential microbial regulatory axis.

## Introduction

The prenatal environment critically influences the developing foetal immune system, with lasting implications for immune and metabolic function, and long-term health outcomes (Durack and Lynch, 2019; Euclydes et al., 2024). Foetal immune development is essential for preparing the neonate for exposure to a microbial environment and establishing lifelong health. Consequently, the timing and nature of prenatal exposures are pivotal in determining their impact on health and development. Although the neonatal innate immune system can immediately respond to pathogens and environmental stimuli at birth, its full maturation requires coordination with the microbiota and other environmental factors (Torow et al., 2017; Hornef and Torow, 2020; Park et al., 2020; Kalbermatter et al., 2021). Maternal-foetal immune crosstalk involves the transfer of maternal antibodies, ingested allergens, maternal bacteria, microbiota-derived metabolites, immune cells, and cytokines across the placenta and is also facilitated by foetal swallowing (Underwood et al., 2005; Burgener and Schroder, 2021; Fernandes and Lim, 2024; Moreno et al., 2024).

The debate regarding uterine sterility during pregnancy and the presence of bacteria in uncomplicated full-term pregnancies persists (Lim et al., 2018; Liu et al., 2020; Blaser et al., 2021; Wang et al., 2022; Banchi et al., 2024). Nevertheless, several studies have detected microbial presence and bacterial DNA in foetal membranes, the placenta, amniotic fluid (AF), and umbilical cord blood prenatally (Rautava et al., 2012; Stout et al., 2013; Aagaard et al., 2014; Collado et al., 2016; Bassols et al., 2016; Vytla et al., 2016; Stinson et al., 2017; Zhu et al., 2018; Stinson et al., 2019a; Younge et al., 2019; Stinson et al., 2020; Wang et al., 2021; Campisciano et al., 2023; Zhong et al., 2024). Both culture-dependent and culture-independent methods have also identified bacterial populations initiating foetal intestinal colonization *in utero* without apparent adverse effects on pregnancy or infant health (Rackaityte et al., 2020; Li et al., 2020; Mishra et al., 2021).

Although the phenomenon of microbial colonization in the placental foetal compartment is not yet fully understood, several mechanisms have been proposed to explain how microbes may reach this environment. This includes ascending pathways originating from the vagina (Dera et al., 2024), active sampling of the intestinal lumen by maternal dendritic cells (Schoenmakers et al., 2018), and hematogenous dissemination to the placenta (Loughran and Tuomanen, 2016). Furthermore, bacterial translocation via the bloodstream is enhanced during pregnancy and lactation due to weakened mucosal barriers, facilitating bacterial entry into circulation and placental seeding (Romano-Keeler and Weitkamp, 2015; Yu et al., 2021).

Cultivation-dependent studies likely underestimate microbial prevalence due to the challenges of culturing anaerobic bacteria or those requiring specific, unidentified nutrients (Bonnet et al., 2020; Banchi et al., 2024). Conversely, Next-Generation Sequencing (NGS) technologies face limitations in microbial community profiling due to the difficulty in distinguishing authentic signals from inherent background contamination in low-biomass samples (Biesbroek et al., 2012; Salter et al., 2014; Glassing et al., 2016; de Goffau et al., 2018; Stinson et al., 2019b; Eisenhofer et al., 2019; Olomu et al., 2020; Nearing et al., 2021).

To minimise heterogeneity across studies and improve their comparability for drawing meaningful conclusions, numerous authors emphasise the need for enhancements in sample collection and processing, as well as the establishment of standardised guidelines for both culture-based and DNA-based sequencing studies (Silverstein and Mysorekar, 2021; Banchi et al., 2024). To overcome the limitations above mentioned in previous studies, we explored microbial colonization in meticulously collected AF samples from two distinct cohorts of women (in their second and third trimesters of pregnancy) from different hospitals, with none having initiated labour and without clinical and analytical suspicion of AF infection. Our approach integrates both culturing techniques and various NGS technologies. Notably, this dual-methodology study in healthy women across two gestational periods, considers the intrinsic differences in the metabolic environment of AF samples and includes an investigation of antimicrobial peptides (AMPs). This approach offers a novel perspective compared to previous research on the foetal microbiome. Within this framework, and based on the results of this extensive study, we underscore the significance of microbial presence in the intrauterine environment, suggesting the potential for a prenatal microbial regulatory axis.

## Results

### Optimised procedures for AF microbial culture analysis

In this study, we improved and fine-tuned our laboratory procedures to maximise the growth and isolation of microorganisms from AF samples. Specifically, we optimised procedures for microbial culture in plates using a subset of randomly selected AF samples (n=10), selecting the most suitable culture media, adjusting incubation times, and refining pre-enrichment conditions. Additionally, we developed a rigorous protocol to minimise contamination from reagents and handling procedures (see Materials and Methods).

We verified the results of this optimization by colony counting on plates. Among the various concentration and centrifugation conditions tested, a 5-fold concentration of AF combined with centrifugation at 9,500 × g for 12 minutes exclusively yielded colony growth.

Four media types were tested with AF samples (n=5) under both aerobic and anaerobic conditions. Colonies grew on all media except MC agar, which was subsequently excluded. Most aerobic colonies developed within 48 to 96 hours, though some required up to 5 days. The pre-enrichment in BHI medium proved crucial for detecting slow-growing or initially undetectable microorganisms. Finally, no microbial growth was observed on plates from any of the negative control samples used in the study.

### Profiling of culturable bacteria in amniotic fluid samples

In this study, we investigated the presence of microorganisms in AF samples collected during different stages of pregnancy. Samples were obtained via amniocentesis (n=127) and elective caesarean section (n=27). To prevent potential distortion of results, 12 AF samples (five from amniocentesis and seven from elective caesarean sections) were excluded due to blood contamination or insufficient volume (**Fig. 1**).

**Figure 1.**
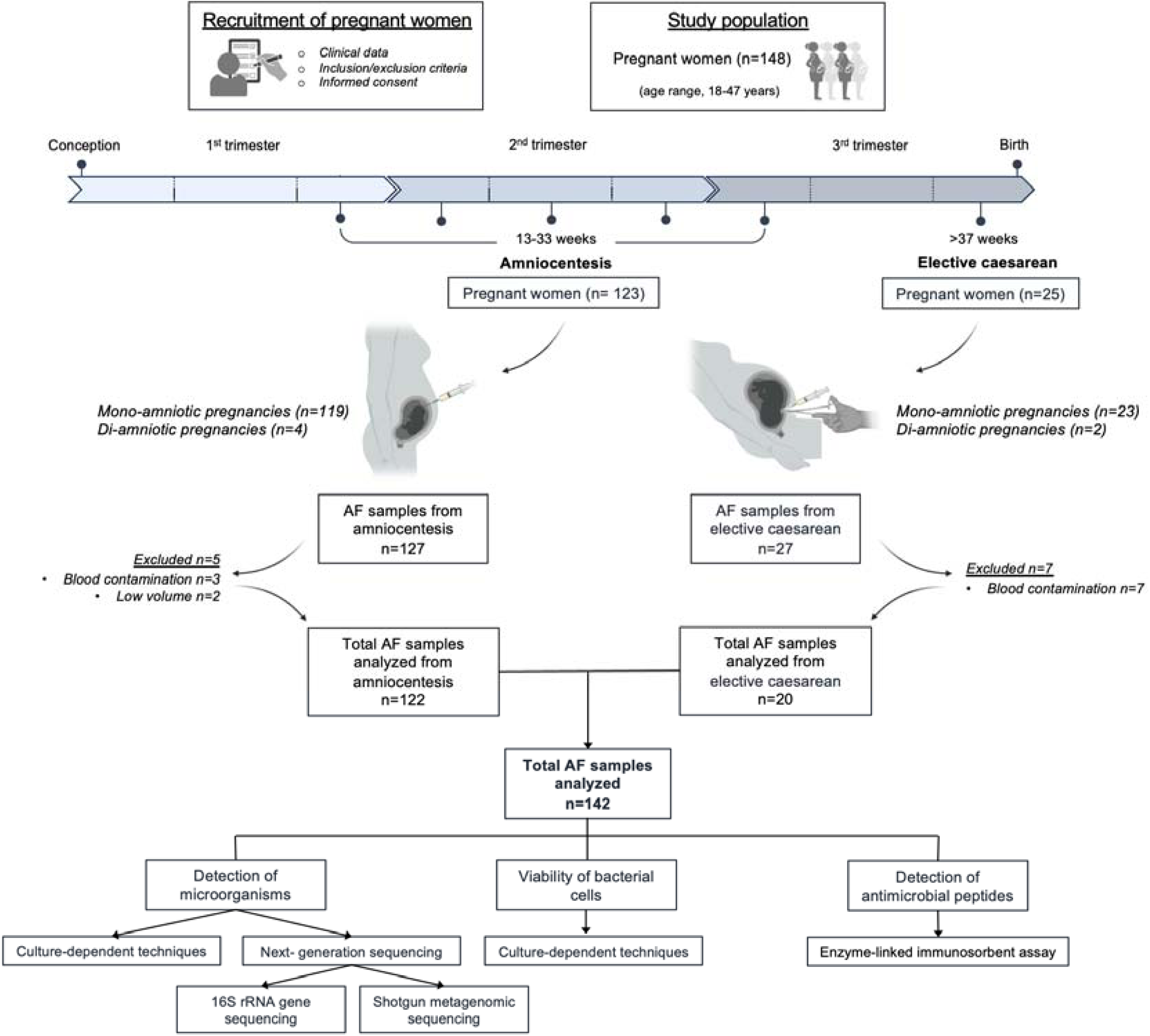
Flowchart of study design and pregnancy outcomes. AF, amniotic fluid.

AF samples (n=142) were concentrated and subsequently cultured under optimised procedures, following the standardised protocol, encompassing both aerobic and anaerobic environments. Out of the 142 AF samples analysed, 33.1% contained culturable microorganisms. Specifically, 29.5% (36/122) of amniocentesis samples and 55.0% (11/20) of caesarean section samples tested positive. To identify bacteria recovered from AF samples using culture from both gestational periods, the 16S rRNA gene was PCR-amplified. Species in the Venn diagram (**Fig. 2A**) represent the closest identified bacterial matches (≥99% homology). Microorganisms from the phyla Actinobacteria, Firmicutes and Proteobacteria were identified. Interestingly, species such as *Bacillus subtilis*, *Cutibacterium acnes*, *Micrococcus luteus*, and *Staphylococcus epidermidis* were present in AF samples from both gestational periods. Regarding abundance, *C. acnes*, *M. luteus*, and *S. epidermidis* predominated in second trimester samples, whereas *Staphylococcus lugdunensis*, *B. subtilis*, and *S. epidermidis* were most prevalent in term deliveries.

**Figure 2.**
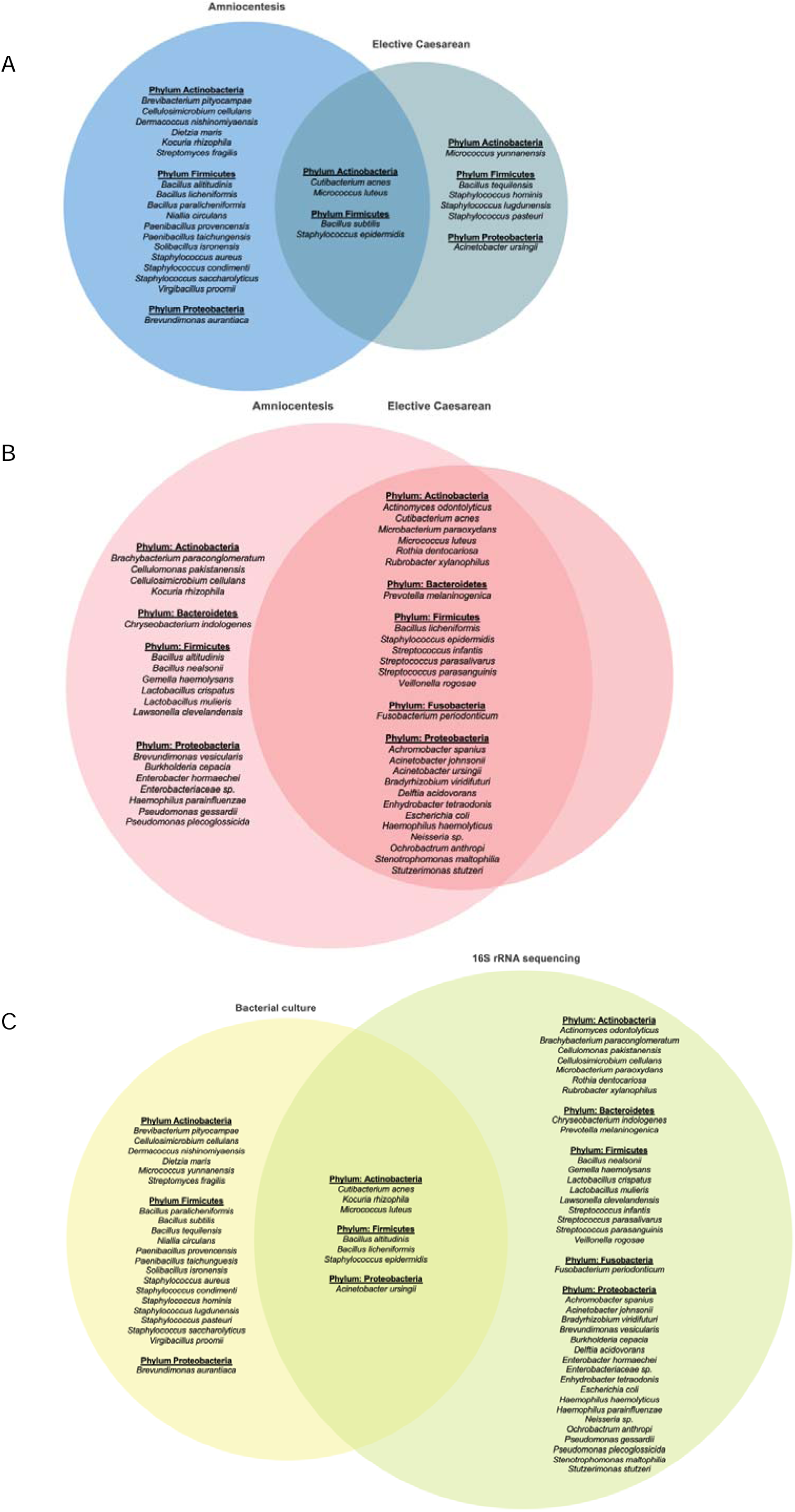
Venn diagram. **(A)** Cultivable bacterial species isolated from AF samples from amniocentesis and elective caesarean sections. **(B)** Bacterial species identified by 16S rRNA sequencing from AF samples from amniocentesis and elective caesarean sections. **(C)** Cultivable bacterial species isolated and bacterial species identified by 16S rRNA sequencing from AF samples from amniocentesis and elective caesarean sections. AF, amniotic fluid.

Given previous studies suggesting the potential for microbiological contamination in the analysis of AF samples, we compared the 16S rRNA gene similarity of individual isolates. A thorough phylogenetic analysis (**Fig. S1**) was performed on AF samples by both amniocentesis and caesarean sections, categorised by the hospital where each sample was collected. Except for *B. subtilis*, bacterial species were isolated from women across all three participating hospitals. The phylogenetic analysis revealed several patterns within the studied species. Isolates from caesarean sections often clustered with those from amniocentesis, indicating no distinct grouping based on gestational age or hospital of origin. Across all species studied, isolates formed clear, distinct clusters and branches, with no correlation found related to the sample source.

Next, we assessed microbial colonization in AF samples by counting isolated colonies. Of the culture-positive amniocentesis plates, 96.7% yielded a single colony. In contrast, caesarean section samples showed more varied patterns: five samples had a single colony, while 54.54% exhibited multiple colonies, ranging from 2 to 23 per plate. In total, 147 microbial colonies were identified in AF samples: 82 from amniocentesis (47 aerobic and 35 anaerobic) and 65 from caesarean section (12 aerobic, and 53 anaerobic). Colony counts differed significantly between aerobic and anaerobic conditions (*p*=0.0005), with caesarean section samples showing a higher number of colonies under anaerobic conditions. No significant differences in colony counts were observed between BHI and CB media.

The absolute abundance of culturable bacterial species per AF sample was also analysed, providing an overview of the microbial community diversity in AF samples obtained through amniocentesis and elective caesarean section. In 47.22% (17/36) of all culture-positive amniocentesis samples, a single species was isolated. Another 30.56% (11/36) contained two species, 8.33% (3/36) had three species, and 13.89% contained more than three. In contrast, among caesarean section samples, 45.45% (5/11) had a single species, while 36.36% (5/11) contained two species and 18.18% (2/11) had more than 3 species. One elective caesarean sample, in which six microbial species were identified, showed the highest number of colonies for each of these bacterial species.

### The impact of the amniotic cavity on bacterial viability: assessing growth in AF samples

To investigate the intrinsic capacity of AF to support bacterial viability and proliferation even under forced, non-physiological conditions atypical of the *in utero* environment, we designed a series of experiments using AF samples confirmed to lack detectable cultivable microorganisms using our optimised culture methods.

Initially, seven AF samples (AF_18, AF_36, AF_37, AF_41, AF_42, AF_43 and AF_44), verified to be free of detectable cultivable microorganisms, were incubated in sterile microcentrifuge tubes under both aerobic and anaerobic conditions. Growth was assessed via absorbance and measurements recorded. A parallel water-only and culture medium negative controls were incubated in identical tubes and conditions. We observed an increase in absorbance in these AF samples after 96 hours, suggesting the presence of microorganisms at low or dormant levels, which were able to proliferate to detectable concentrations due to prolonged incubation and the experimental conditions (**Fig. S2A**). The subsequent plating of the enriched AF sample on solid media led to the isolation of *Amycolatopsis pretoriensis* and *Cellulosimicrobium funkei*, underscoring the value of this approach for discovering low-abundance or slow-growing microbes in AF.

In a subsequent experiment, we used AF samples (AF_31, AF_82, AF_83, AF_84 and AF_133) confirmed to be free of culturable bacteria to assess their ability to support the growth of bacterial isolates. For this, four of the most isolated species from the previous solid medium culture assay (*B. subtilis*, *C. acnes*, *M. luteus*, and *S. epidermidis*) were individually inoculated into AF samples using sterile microcentrifuge tubes. Negative water controls were also established. Our results indicated substantial variations in microbial growth among the tested species, contingent on the specific AF sample serving as the culture medium (**Fig. S2B**). Furthermore, differential growth patterns were evident among the various bacterial species within the same AF sample. Of the four species analysed, *C. acnes* consistently demonstrated the highest growth, whereas *S. epidermidis* displayed the lowest proliferative capacity across all AF samples.

### Microbial composition identified by 16S rRNA gene sequencing

Although metagenomic studies typically yield valuable insights, their application to AF samples critically depends on rigorous controls and careful interpretation. This is essential due to the significant challenges posed by the low microbial biomass niche and the persistent risk of environmental contamination and kit-derived contamination (“kitome”).

In this study we first assessed the influence of DNA extraction kits on bacterial DNA detection in AF samples by comparing two extraction protocols, evaluating levels of contaminants, and distinguishing true microbial signals from background noise using negative controls.

Genomic DNA was extracted from 18 randomly selected AF samples and four blank extraction controls, using both the PowerFecal Pro (PFP) and MagAttract Microbial DNA (MA) kits. Full-length 16S rRNA gene sequencing was then performed. This comparative approach enabled us to evaluate the performance of each kit in terms of operational taxonomic unit (OTU) recovery and the extent of kitome contamination (Salter et al., 2014; Glassing et al., 2016; Stinson et al., 2019b; Velásquez-Mejía et al., 2018). Blank extraction controls were included in each extraction batch.

All negative controls were quantified using a high-sensitivity Qubit assay and fell below the detection limit, indicating minimal contamination during DNA extraction. As shown in **Table 1**, the PFP kit produced a lower average read count in negative controls compared to the MA kit, suggesting reduced background contamination. In the AF samples, reads corresponding to several bacterial taxa were detected exclusively in the AF samples and not in the negative controls. The only two species shared between the AF samples and negative controls were *C. acnes* and *S. epidermidis*. Notably, the PFP protocol yielded a higher average read depth in AF samples, indicating improved sensitivity for detecting microbial DNA. To evaluate potential contamination introduced during PCR amplification, four negative PCR controls were included, which yielded an average of 140.75 reads. Only reads corresponding to three bacterial species were detected in these controls: *C. acnes*, *Exiguobacterium profundum*, and *S. epidermidis*, although not a significant level in relation to the overall reads obtained; among these, only *C. acnes* and *S. epidermidis* were also identified in AF samples (**Table 2**).

**Table 1.** Comparison of bacterial taxa identified by 16S rRNA gene sequencing in negative controls and random AF samples using two, PFP and MA, extraction kits. AF, amniotic fluid; MA; Mag Attract; PFP, PowerFecal Pro.

| Extraction kit | Negative controls (n=4) |  | Random AF samples (n=18) |  |
| --- | --- | --- | --- | --- |
|  | OTUs | Reading average | OTUs | Reading average |
| **PFP** | *Alkalihalobacillus alcalophilus*<br>*Anaerobacillus isosaccharinicus*<br>***Cutibacterium acnes***<br>*Evansella tamaricis*<br>*Methylophilus medardicus*<br>*Pseudomonas gessardii*<br>***Staphylococcus epidermidis*** | 1,763.50 | *Acinetobacter johnsonii*<br>*Bacillus licheniformis*<br>*Bradyrhizobium viridifuturi*<br>*Brachybacterium paraconglomeratum*<br>*Burkholderia cepacia*<br>*Cellulosimicrobium cellulans*<br>*Cellulomonas pakistanensis*<br>***Cutibacterium acnes***<br>*Enhydrobacter tetraodonis*<br>*Escherichia coli*<br>*Haemophilus parainfluenzae*<br>*Kocuria rhizophila*<br>*Microbacterium paraoxydans*<br>*Micrococcus luteus*<br>*Neisseria sp.*<br>*Pseudomonas plecoglossicida*<br>*Rubrobacter xylanophilus*<br>***Staphylococcus epidermidis***<br>*Stenotrophomonas maltophilia*<br>*Streptococcus infantis*<br>*Streptococcus parasalivarius*<br>*Streptococcus parasanguinis*<br>*Veillonella rogosae* | 50,915.72 |
| **MA** | *Acinetobacter indicus*<br>*Alkaliphilus transcaalensis*<br>*Chryseomicrobium amylolyticum*<br>***Cutibacterium acnes***<br>*Enterobacteriaceae bacterium*<br>*Erwinia endophytica*<br>*Exiguobacterium profundum*<br>*Pseudomonas gessardii*<br>***Staphylococcus epidermidis*** | 10,969.00 | *Bacillus licheniformis*<br>*Bradyrhizobium viridifuturi*<br>***Cutibacterium acnes***<br>*Escherichia coli*<br>*Haemophilus parainfluenzae*<br>*Kocuria rhizophila*<br>*Micrococcus luteus*<br>*Neisseria sp.*<br>*Pseudomonas plecoglossicida*<br>*Rubrobacter xylanophilus*<br>***Staphylococcus epidermidis***<br>*Stenotrophomonas maltophilia*<br>*Streptococcus parasanguinis*<br>*Veillonella rogosae* | 33,149.22 |

**Table 2.**
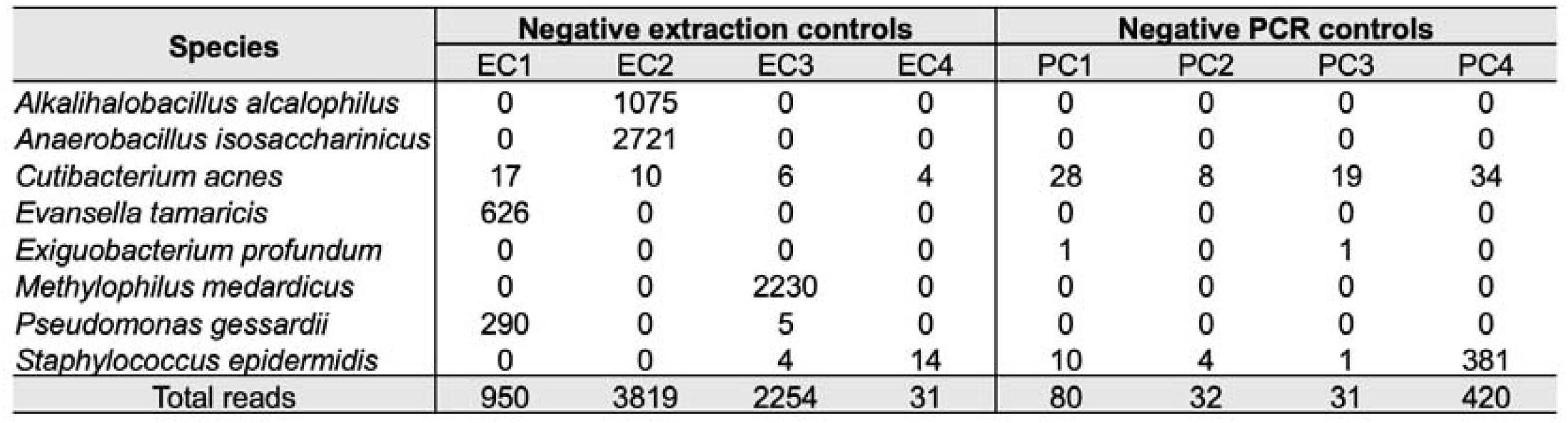
16S rRNA gene sequence reads obtained from negative extraction controls (EC1-4) using the PFP kit, and from negative PCR controls (PC1-4). PFP, PowerFecal Pro.

| Species | Negative extraction controls |  |  |  | Negative PCR controls |  |  |  |
| --- | --- | --- | --- | --- | --- | --- | --- | --- |
|  | EC1 | EC2 | EC3 | EC4 | PC1 | PC2 | PC3 | PC4 |
| *Alkalihalobacillus alcalophilus* | 0 | 1075 | 0 | 0 | 0 | 0 | 0 | 0 |
| *Anaerobacillus isosaccharinicus* | 0 | 2721 | 0 | 0 | 0 | 0 | 0 | 0 |
| *Cutibacterium acnes* | 17 | 10 | 6 | 4 | 28 | 8 | 19 | 34 |
| *Evansella tamaricis* | 626 | 0 | 0 | 0 | 0 | 0 | 0 | 0 |
| *Exiguobacterium profundum* | 0 | 0 | 0 | 0 | 1 | 0 | 1 | 0 |
| *Methylophilus medardicus* | 0 | 0 | 2230 | 0 | 0 | 0 | 0 | 0 |
| *Pseudomonas gessardii* | 290 | 0 | 5 | 0 | 0 | 0 | 0 | 0 |
| *Staphylococcus epidermidis* | 0 | 0 | 4 | 14 | 10 | 4 | 1 | 381 |
| Total reads | 950 | 3819 | 2254 | 31 | 80 | 32 | 31 | 420 |

After this analysis, an additional 73 AF samples were subjected to sequencing using the PFP kit. To standardize sequencing depth, subsampling was performed to 1,413 reads, based on the smallest library size, excluding negative controls. We obtained an average of 53,043.82 reads per AF sample collected via amniocentesis, corresponding to an average of 5.86 OTUs per sample. Similarly, AF samples collected during elective caesarean sections yielded an average of 34,665.15 reads and 4.38 OTUs per sample. Overall, AF samples exhibited a bacterial community characterised by low abundance and limited diversity. Notably, all bacterial species identified in AF samples collected during caesarean sections were also present in those obtained by amniocentesis (**Fig. 2B**). The sequenced bacterial phyla included Actinobacteria, Bacteroidetes, Firmicutes, Proteobacteria, and Fusobacteria.

To assess the consistency and complementarity between traditional culture-based methods and culture-independent 16S rRNA gene sequencing, we compared the bacterial taxa identified by each approach. As shown in **Fig. 2C**, a total of 65 bacterial taxa were detected across both methods. Of these, seven taxa were identified by both culture and 16S rRNA gene sequencing, and 37 taxa were detected exclusively through sequencing. The taxa shared by both methodologies included representatives from the phyla Actinobacteria (*Kocuria rhizophila*, *C. acnes* and *M. luteus*), Firmicutes (*Bacillus licheniformis*, *Bacillus altitudinis* and *S. epidermidis*), and Proteobacteria (*Acinetobacter ursingii*). Culture-based methods recovered several unique taxa, primarily within the Firmicutes and Actinobacteria phyla. Conversely, 16S rRNA gene sequencing offered broader phylogenetic coverage, detecting additional phyla such as Bacteroidetes (*Chryseobacterium indologenes* and *Prevotella melaninogenica*) and Fusobacteria (*Fusobacterium periodonticum*), which were not identified by culture.

The relative abundance of bacterial species detected in AF samples across different gestational age groups was also analysed (**Fig. 3A**). The microbial profiles exhibited considerable inter-individual variability, with no single taxon predominating consistently across all samples. *C. acnes* and *S. epidermidis* were among the most consistently abundant species throughout all trimesters, corroborating their frequent detection by both culture and sequencing methods. *C. acnes* was present in 84.27% of the samples, while *S. epidermidis* appeared in 76.40%, highlighting their widespread occurrence within the cohort.

**Figure 3.**
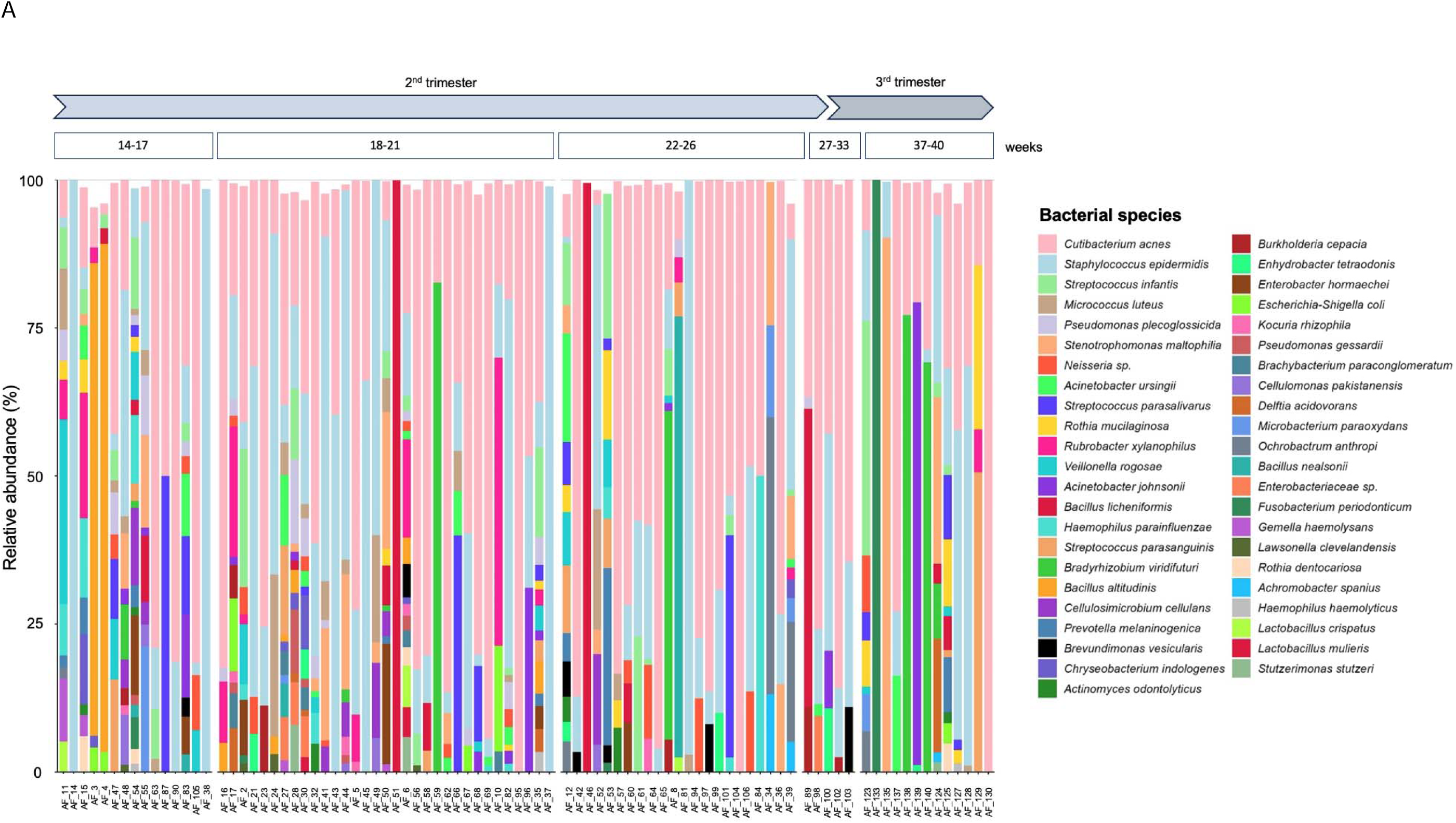
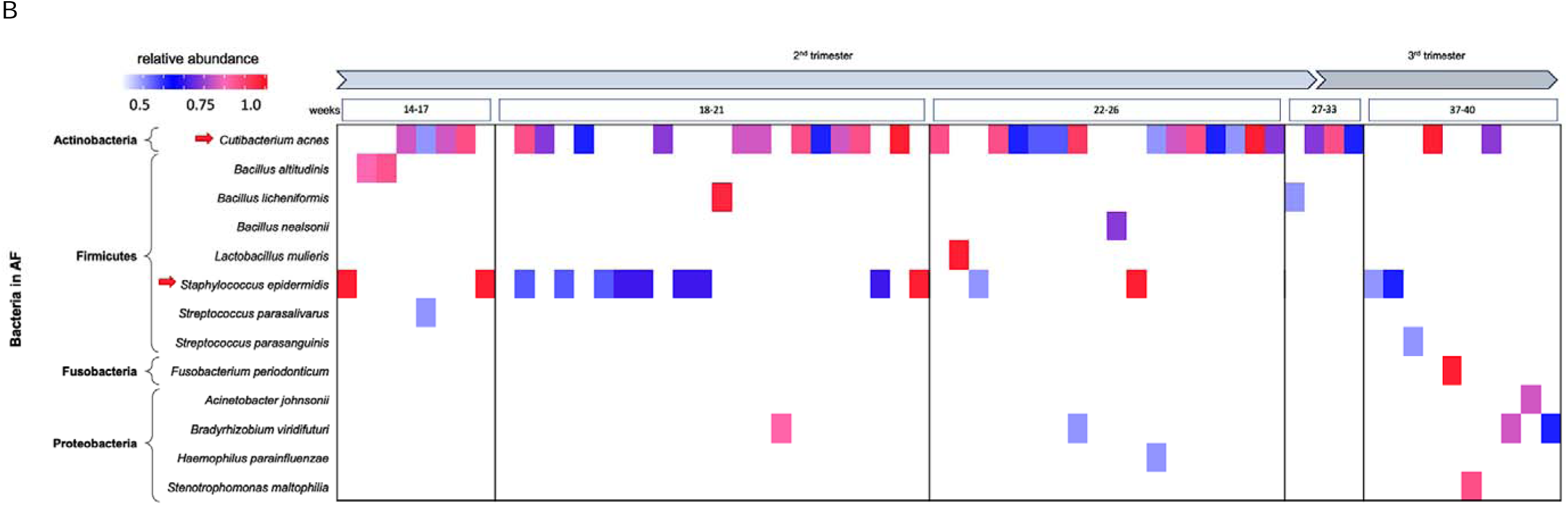
**(A)** Relative abundance (%) of bacterial species detected in AF samples by 16S rRNA gene sequencing, categorised by gestational age and weeks of pregnancy. **(B)** Heatmap of species with >0.5 relative abundance obtained after 16S rRNA gene sequencing analysis. High bacterial relative abundance is represented with color gradation which increases toward red, while low bacterial relative abundance tends toward blue. The arrow indicates the most abundant species identified. AF, amniotic fluid.

To further investigate the composition of the dominant microbial communities, OTUs with a relative abundance greater than 0.5 were stratified by gestational age. In **Fig. 3B**, the heatmap highlights a small subset of taxa that achieve high relative abundance in AF samples, indicating that most detected microorganisms are low in biomass and sporadic in occurrence. Among these, *C. acnes* and *S. epidermidis* stand out as the most prominent taxa, not only due to their widespread presence across all gestational periods but also because they reach high relative abundance values (>0.75) in several samples, particularly between weeks 14-17 and 22-26.

Other taxa, such as *B. licheniformis*, *Lactobacillus mulieris*, and *F. periodonticum*, were detected less consistently, each appearing in only a single sample. However, in those cases, they nearly dominated the microbial community with relative abundance values approaching 1.0, suggesting episodic but pronounced microbial presence in specific individuals.

### Microbial composition identified by shotgun metagenomic sequencing

To complement the taxonomic insights from 16S rRNA gene sequencing, we performed shotgun metagenomic sequencing on five AF samples selected by presence/absence of culturable bacteria. This approach enabled a more comprehensive characterization of microbial communities, offering strain-level resolution. The sequencing results revealed a taxonomically diverse, yet low-biomass microbial community, with notable variability in microbial content across samples.

Shotgun metagenomic sequencing yielded approximately 1.2 to 8.3 million raw reads per AF sample, encompassing both microbial and host reads. A high percentage of these sequences were classified across all samples (ranging from 76.5% to 98%, see **Table 3**). The proportion of microbial reads varied notably, from less than 5% in AF_9 to over 60% in AF_29. Bacterial reads constituted the majority of the microbial content in each sample, with AF_29 and AF_35 showing particularly high percentages (59.33% and 35.19%, respectively).

**Table 3.** Summary of sequencing classification results for five samples analysed by shotgun metagenomics. The table displays the total number of reads, the percentage of classified reads, and the proportions of microbial and bacterial reads. Asterisks (*) indicate samples with the highest relative abundance in each microbial category.

| Samples | Total reads | Percentage (%) |  |  |
| --- | --- | --- | --- | --- |
|  |  | reads classified | microbial reads | bacteria reads |
| AF_12 | 1,975,495 | 97.76 | 7.72 | 0.65 |
| AF_13 | 1,473,926 | 98.03 | 9.32 | 5.51 |
| AF_29 | 8,387,313 | 76.51 | 62.83 | 59.33* |
| AF_9 | 1,261,771 | 97.63 | 4.93 | 1.30 |
| AF_35 | 2,681,675 | 85.39 | 38.98 | 35.19* |

Among the detected taxa, *Phyllobacterium* was the predominant genus in two samples from which high-quality metagenome-assembled genomes (MAGs) were recovered. These MAGs were over 90% complete with low contamination. *Phyllobacterium* sp. T1293, *Phyllobacterium* sp. 628, and *Phyllobacterium zundukense* were particularly abundant species, suggesting that multiple members of this genus are well-adapted to the mildly oxygenated intra-amniotic environment. Other bacterial genera were identified at lower abundance, indicating a microbial community dominated by a few key species (**Fig. 4**).

**Figure 4.**
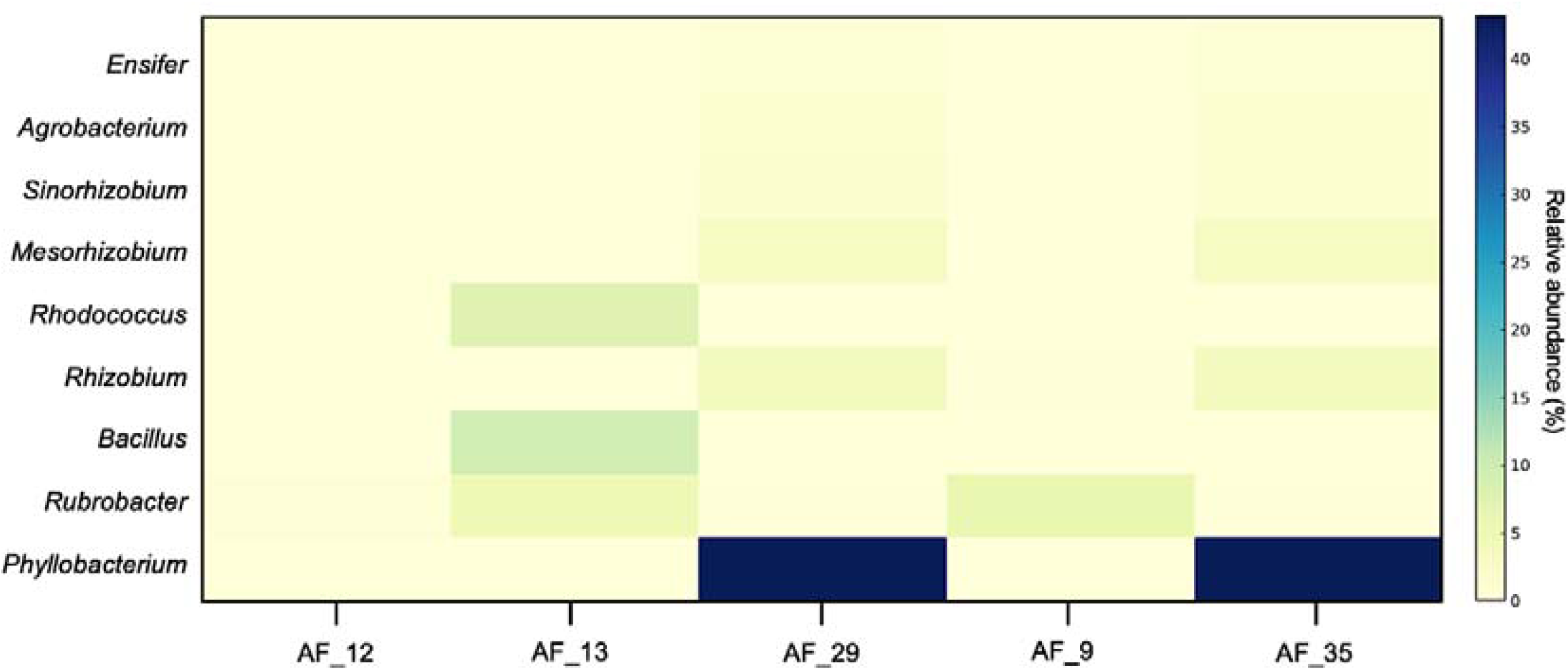
Relative abundance (%) of dominant bacterial genera detected by shotgun metagenomics across samples. Only genera with >0.5% relative abundance are shown. Color intensity reflects the proportion of taxonomically classified reads assigned to each genus based on shotgun sequencing data.

### Impact of different clinical and procedural aspects on the presence of microorganisms in AF

To further investigate the overall microbial community structure across different clinical and procedural variables, we performed a beta diversity analysis using non-metric multidimensional scaling (NMDS) on our 16S rRNA gene data. As shown in **Fig. 5**, the ordination plots illustrate sample clustering according to (A) gestational period (amniocentesis *versus* elective caesarean sections) (R^2^=0.01421; *p*=0.1039), (B) sample collection centre (R^2^=0.01275; *p*=0.2796), and (C) foetal condition (normal, chromosomopathy, or structural malformation) (R^2^=0.0095; *p*=0.6262). Although some visual grouping can be observed in the NMDS plots, pairwise PERMANOVA analyses did not reveal statistically significant differences between any of the groups (*p*<0.05), indicating that the global microbial composition shows substantial inter-sample variability across these conditions. No significant differences in microbial composition were detected when samples were stratified by weeks of gestational age (R^2^=0.1831, *p*=0.8937), assisted *versus* spontaneous conception (R^2^=0.02229, *p*=0.0767) or pregnancy outcome (R^2^=0.00996, *p*=0.8202).

**Figure 5.**
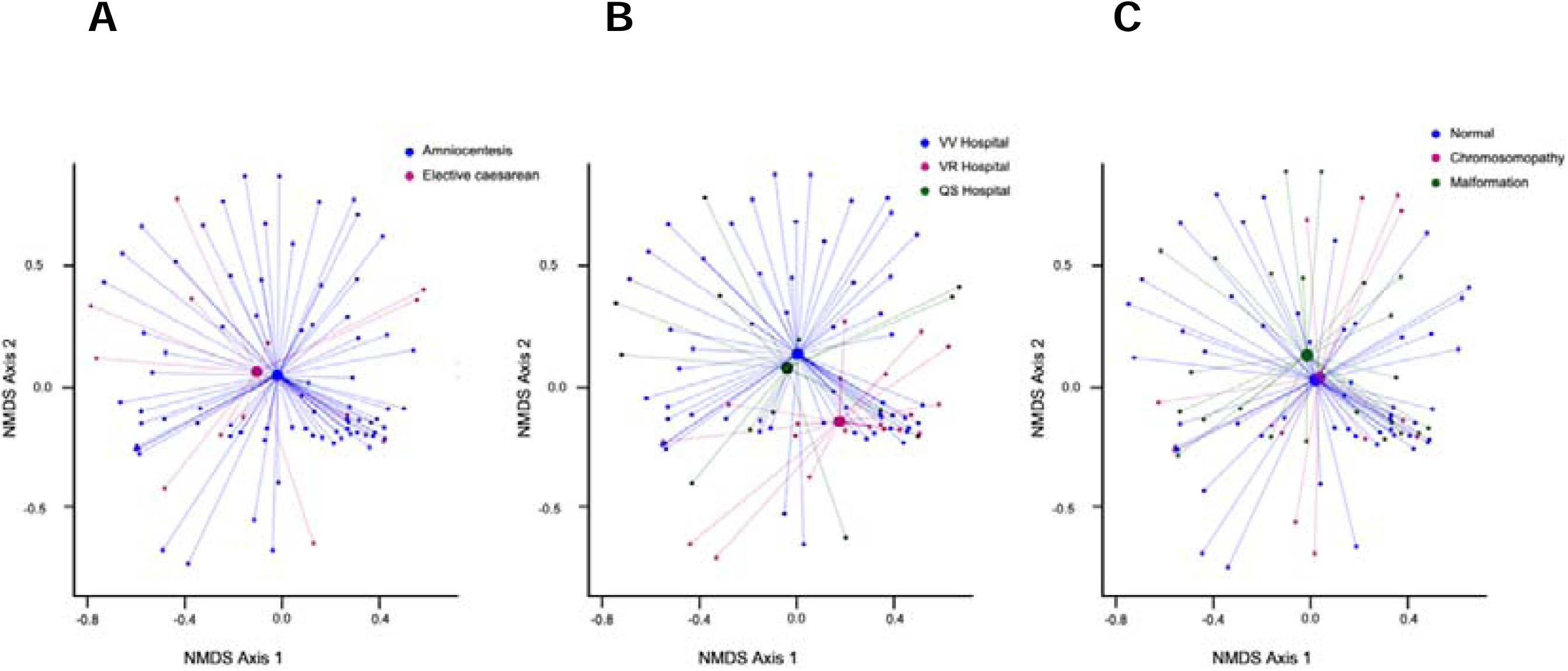
Beta diversity analysis by NMDS and PERMANOVA. Samples grouped according to: **(A)** gestational period (R^2^=0.01421; *p*=0.1939), **(B)** sample collection center (R^2^=0.01275; *p*=0.2796), and **(C)** foetal status features: normal, malformations, and chromosomopathies (R^2^=0.0095; *p*=0.6262). Clustering patterns represent differences in microbial community composition among groups. NMDS, non-metric multidimensional scaling. \**p*<0.05

In contrast, the analysis of results from culturable bacteria in AF samples revealed a significant association between isolated bacteria and the type of reproductive treatment (n=10, assisted conception; n=90, spontaneous conception; *p*=0.028). However, culture positivity showed no significant relationship with gestational age (*p*=0.9017), maternal age (*p*=0.703), foetal status (*p*=0.1692), or pregnancy outcome (*p*=0.7607). These findings indicate that, within this cohort, the presence of culturable bacteria is not dependent on most maternal or foetal clinical characteristics.

### Microbial intra-individual variability in diamniotic twin AF

To evaluate the microbial composition of diamniotic AF samples, we identified bacterial species in paired amniotic sacs from three individuals using both culture-based methods and 16S rRNA gene sequencing. In the AV_3 amniocentesis case, 16S rRNA gene sequencing identified five bacterial taxa in each sac, with shared species including *B. altitudinis*, *C. acnes*, and *Escherichia coli*. Notably, culture methods detected *Solibacillus isronensis* exclusively in sac 2, while sac 1 yielded no culturable bacteria. These findings suggest a similar microbial community at the sequencing level, but limited overlap in culture results between sacs. For the AV_60 amniocentesis case, *S. epidermidis* was consistently detected by both culture and sequencing in sac 1. *C. acnes* was also found in both sacs by at least one detection method. Sac 2 displayed greater microbial diversity via 16S rRNA gene methods, identifying six species compared to two in sac 1. This indicates partial overlap alongside distinct differences in microbial content between sacs. In the EC_5 elective caesarean section case, *S. epidermidis* was the sole species recovered by culture (sac 1) and was also detected by 16S rRNA gene sequencing in both sacs. However, sequencing identified a broader range of taxa in sac 1 (*Haemophilus haemolyticus*, *Rothia mucilaginosa* and *Streptococcus parasalivarius*), whereas sac 2 showed fewer taxa and no culture-positive results.

### Analysis of AMP in AF: association with gestational stages and microbial detection

To further investigate the potential relationship between AMPs and the presence of microorganisms in AF during pregnancy, we quantified the levels of α-defensins (HNPs 1–3), β-defensins (HBD-1, HBD-2, HBD-3), and cathelicidins (LL-37) in randomly selected AF samples (**Table S1**). Of these, 7 were obtained via amniocentesis and 4 via elective caesarean sections, and all belonged to either healthy foetuses or those with abnormalities. Among the amniocentesis samples, 3 tested positive (AF_87, AF_52 and AF_53) for culturable microorganisms, while 4 (AF_83, AF_84, AF_39 and AF_68) were negative. In the elective caesarean section group, microbial cultures were positive in half of the samples (AF_124 and AF_129) and negative (AF_133 and AF_140) in the other half. Regarding the metagenomic characterization of the analysed samples for AMPs, 16S rRNA gene sequencing exhibited detectable microbial DNA with taxonomic profiles exceeding a predefined relative abundance threshold of 0.05%.

HBD-1, HBD-2, HBD-3, and HNP 1-3 levels did not exhibit significant differences across trimesters. However, LL-37 concentrations were significantly higher in amniocentesis samples compared to those from elective caesarean sections (*p*=0.042). Notably, LL-37 was consistently the most abundant AMP detected in AF samples (**Table S1**).

Regarding associations between AMP levels and microbial presence, most peptides did not exhibit statistically significant differences when comparing samples with and without cultivable microorganisms. However, a significant difference was observed for HBD-1 (*p*=0.018), with lower concentrations in samples testing positive for culturable bacteria. Further, when grouping samples by total relative microbial abundance (threshold: >0.05%), HBD-1 concentrations were significantly lower in samples with higher microbial loads (*p*=0.032). No statistically significant differences were found for the remaining peptides under this comparison.

Stratifying samples by the presence of specific genera revealed additional significant findings (**Table S1**). Samples positive for *Staphylococcus* in culture showed significantly reduced concentrations of HBD-1 (*p*<0.001), HBD-3 (*p*=0.024), and LL-37 (*p*=0.049). No significant associations were found between peptide levels and the presence of other cultured genera such as *Bacillus, Cutibacterium, Micrococcus, Paenibacillus,* or *Brevundimonas*. No significant associations were found between AMP levels and bacterial species detected through 16S rRNA gene sequencing.

## Discussion

The microbial composition of AF remains a subject of intense scientific inquiry, particularly in relation to the long-standing debate over sterility *versus* non-sterility (Aagaard et al., 2014; Collado et al., 2016; Lim et al., 2018; Zhu et al., 2018; Stinson et al., 2019a; Younge et al., 2019; Liu et al., 2020; Stinson et al., 2020; Wang et al., 2021; Wang et al., 2022; Campisciano et al., 2023; Banchi et al., 2024; Zhong et al., 2024). Emerging evidence increasingly challenges the traditional paradigm of a sterile womb, suggesting instead the presence of a diverse and dynamic microbial landscape during pregnancy. Advances in NGS have increasingly supported the presence of a low-biomass microbial community in AF, even in the absence of clinical infection (Collado et al., 2016; Willyard, 2018; Stinson et al., 2020). Thus, the major question appears to no longer be whether the prenatal environment is sterile or non-sterile. This study might suggest a possible microbial balance in the prenatal environment, likely providing insights into the relationship between AMPs and early microbial landscape, including the novel concept of a potential prenatal microbial regulatory axis.

We analysed different factors that may contribute to maintaining balanced microbial colonization in the foetus. For that, to characterise the microbial profiles, we optimised culture methodologies alongside 16S rRNA gene and shotgun metagenomic sequencing to achieve a comprehensive characterization of microbial profiles in AF samples collected at various gestational stages. Our approach overcomes limitations of previous studies (Park et al., 2023) by analysing microbial colonization in rigorously collected AF samples from two independent cohorts of women at different hospitals. Importantly, this study accounts for intrinsic differences in foetal status and rigorous contamination control. Secondly, we explored the presence of AMPs in AF, offering new potential insights into their regulatory functions in shaping the prenatal microbial landscape.

Culturable microorganisms were detected in approximately one-third of all AF samples. Notably, 29.51% of amniocentesis-derived samples and 55% of those obtained during caesarean sections yielded positive culture results. These findings are consistent with previous reports suggesting that AF is not entirely sterile and may harbour microorganisms even in uncomplicated pregnancies (Collado et al., 2016; Mishra et al., 2021). Methodological optimization was critical for enhancing the viability of culturable microorganisms and enabled the detection of slow-growing or fastidious organisms typically undetectable under standard microbiological conditions. This protocol minimised bacterial lysis and the release of intracellular contents, thereby ensuring successful subsequent bacterial growth on culture plates. The predominant phyla identified included Actinobacteria, Firmicutes, and Proteobacteria. Interestingly, *B. subtilis*, *C. acnes*, *M. luteus*, and *S. epidermidis* were consistently found across both second trimester (amniocentesis) and term (elective caesarean section) samples. Notably, a greater number of colonies were isolated from caesarean section-derived samples under anaerobic incubation, suggesting a higher prevalence or viability of facultative anaerobes in these cases. These findings underscore the critical importance of optimised culturing techniques for heterogeneous AF samples. The microbial profiles observed in our study support the hypothesis that fluctuating oxygen levels influence the early microbial landscape, even prior to birth. The presence of both aerobic and anaerobic microorganisms in AF samples suggests that the intra-amniotic environment may exhibit dynamic or microaerophilic oxygen conditions, thereby facilitating the establishment of metabolically diverse microbial communities *in utero*. Although direct experimental evidence regarding foetal gut oxygenation remains limited, it has been proposed that the foetal gut represents a mildly oxygenated (aerobic) niche that transitions to anaerobic conditions following birth (Penders et al., 2006; Gosalbes et al., 2013; Pannaraj et al., 2017).

The AF microbial composition identified by 16S rRNA gene sequencing showed a low abundance and low diversity bacterial community. Notably, all bacterial species identified in elective caesarean section samples were also found in those obtained via amniocentesis. The predominant bacterial phyla sequenced included Actinobacteria, Bacteroidetes, Firmicutes, Proteobacteria, and Fusobacteria. These findings underscore the complementary nature of culture-dependent and culture-independent techniques in characterizing AF bacteria, with sequencing revealing a more diverse and phylogenetically rich bacterial community. However, it is important to consider that some of the species detected through 16S rRNA analysis may not be viable, which introduces limitations when interpreting the functional relevance of results obtained from culture-independent methods or cell-free forms (Vernon et al., 2002). Additionally, it cannot be ruled out that bacterial cells are present at concentrations too low to be detected by culturing, even after sample concentration (Liu et al., 2022). Moreover, specific nutritional and/or environmental requirements of the bacteria, may not have been provided by the culture media and conditions used. Circulating cell-free bacterial DNA and peptides may also transfer between mother and foetus, possibly via the umbilical cord, as previously proposed (Lo et al., 1997; Rodríguez et al., 2015). This hypothesis aligns with reports of microbial DNA in placental and amniotic compartments and supports a role for maternal-foetal molecular exchange in shaping the prenatal immune environment (Wang et al., 2021; Xie et al., 2025). The occurrence of immunologically active microbial fragments may contribute to foetal immune priming, even in the absence of live microorganisms. The presence of typical vaginal commensals such as *Lactobacillus* alongside other taxa is not unexpected, given the anatomical proximity and the potential for maternal microbial transmission (Lehtoranta et al., 2022; Campisciano et al., 2023). Their detection may reflect physiological maternal-foetal microbial exchange rather than contamination, particularly when supported by rigorous controls and multiple lines of evidence. In our study, *Lactobacillus* was detected in only 5 out of 91 samples, with just one sample exhibiting a relative abundance greater than 0.5, reaching 0.99. This suggests that although *Lactobacillus* may occasionally be present in the amniotic environment, it is not a predominant component in most samples, highlighting potential differences in microbial transfer dynamics or selective pressures across pregnancies.

The application of 16S rRNA gene sequencing broadened the scope of microbial detection, revealing additional phyla such as Bacteroidetes and Fusobacteria that were not recovered through culture-based methods. These findings reinforce the concept that culture and sequencing offer complementary perspectives, particularly in low-biomass environments. Nonetheless, the overlap between taxa identified by culture and those detected via sequencing was limited to seven, highlighting methodological biases and intrinsic differences in sensitivity and taxonomic resolution between the two approaches.

Among the most frequently isolated taxa were *C. acnes* and *S. epidermidis*, found consistently across gestational periods. While both are commonly regarded as skin commensals and potential contaminants, their repeated detection across multiple samples, clinical sites, and sequencing platforms combined with low read counts in negative controls supports their possible biological relevance in AF. Specifically, both species were identified in the AF samples analysed as well as in the negative controls; however, the number of reads from the negative controls was notably lower (15,75 and 51,75 *C. acnes* and *S. epidermidis*, respectively compared to the mean total reads of 53,043.82 found in AF samples analysed). Their successful cultivation from AF samples suggests that they may not be the result of contamination also supported by the fact that the culture negative controls showed no growth.

Our investigation into the dominant microbial communities in AF revealed no clear trend of microbial abundance or dominance with advancing gestational age. This finding supports the hypothesis of a transient, low-diversity bacterial presence within the amniotic environment. Ultimately, these results reinforce the concept that microbial presence in AF is highly variable between individuals and generally limited to a few dominant taxa per sample, suggesting episodic microbial exposure rather than stable colonization.

Our shotgun metagenomic sequencing analysis provided complementary taxonomic information and enabled higher resolution for certain taxa, particularly concerning the *Phyllobacterium* spp., from which high-quality MAGs were recovered. These MAGs were over 90% complete with low contamination. *Phyllobacterium* sp. *T1293*, *Phyllobacterium* sp. *628*, and *Phyllobacterium zundukense* were particularly abundant species, suggesting that multiple members of this genus are well-adapted to the mildly oxygenated intra-amniotic environment. Metabolic reconstruction indicates that *Phyllobacterium* exhibits a versatile metabolic profile. It requires basic inorganic salts (e.g., phosphate, sulfate, Fe²⁺, Mg²⁺, Mn²⁺, Ca²⁺), small amounts of certain amino acids (e.g., L-serine, L-phenylalanine, L-proline), and vitamins such as thiamin. Additionally, it can metabolize a wide range of carbon sources. This low dependency on complex nutritional factors, combined with its ability to thrive under mild aerobic conditions, supports its persistence in this environment and reinforces the idea that its presence is not incidental but likely under detected by other methods.

Our comprehensive analysis of AF microbial communities across diverse clinical and procedural variables did not reveal conclusive insights into the nature of intrauterine microbial presence, specifically regarding its association with these variables. Despite visual trends observed in NMDS ordination plots, beta diversity analysis did not demonstrate statistically significant differences in overall microbial composition across gestational period, sample collection centre, foetal condition, or gestational age. However, a significant association between bacterial isolation and the type of reproductive treatment was observed. This finding could suggest that assisted reproductive technologies may influence the translocation or detectability of culturable bacteria in the amniotic environment. This finding is highly significant as it suggests that the global microbial profile within the amniotic environment in our studied cohort is largely independent of these major clinical and obstetric factors. This challenges the notion that variations in maternal or foetal characteristics directly lead to substantial shifts in the overall microbial landscape of the AF (Younge et al., 2019).

The inclusion of AF samples from twin pregnancies constitutes a novel and significant aspect of our study. To our knowledge, this dual methodological approach remains largely unexplored in the existing literature in twins. Notably, we observed intra-individual variability in microbial profiles. In diamniotic twin pregnancies, where each foetus develops in a distinct amniotic sac within the same uterine environment, microbial discordance between paired sacs was detected, suggesting localised differences in microbial exposure despite shared maternal conditions. While *C. acnes* and *S. epidermidis* were consistently detected in both sacs, unique taxa were identified in only one sac per pair, indicating a lack of microbial homogeneity even within the same intrauterine environment. Furthermore, NMDS analysis revealed that diamniotic twins did not exhibit a more similar microbial composition than randomly selected sample pairs, as previously reported (Park et al., 2023). This highlights intra-individual variability in microbial composition within diamniotic pregnancies and underscores the enhanced sensitivity of 16S rRNA gene sequencing compared to culture methods.

Our final aim was to correlate the intrinsic antibacterial activity in AF with the microbial colonization of each sample by examining AMPs. Prior research shows that AMPs like defensins and cathelicidins typically change concentration throughout gestation (Varrey et al., 2018). We found that LL-37 was consistently the most abundant AMP across all trimesters, with higher concentrations in amniocentesis samples compared to those from elective caesarean sections. This suggests potential differences in AMPs related to the gestational stage.

Furthermore, we observed elevated HBD-1 levels in samples lacking culturable microorganisms, potentially suggesting an antimicrobial function in preserving possible amniotic balance. Similarly, HBD-3 concentrations were significantly higher in samples negative for microbial growth. The significantly reduced concentrations of HBD-1, HBD-3, and LL-37 in *Staphylococcus*-positive AF samples suggest a possible interaction between this bacterium and the host’s antimicrobial defenses. Specifically, these results could indicate a selective suppression or degradation of these key AMP in the presence of certain bacterial taxa. Collectively, these results may support the potent antimicrobial nature of AMPs, demonstrating their effectiveness in inhibiting microbial proliferation within the amniotic environment. However, further assessment in larger sample sizes is needed to confirm these results.

In conclusion, all these findings indicate that viable bacteria and/or their genetic material may reach the prenatal environment before birth. Although the underlying regulatory mechanisms require further investigation, their presence may be shaped by maternal and foetal immune responses. Future research will be essential to elucidate the direct impact of this microbial interaction on foetal health outcomes.

## Material and methods

### Study design and subjects

This study involved samples from 148 pregnant women randomly recruited from the Virgen de Valme University Hospital (Sevilla, Spain), Virgen del Rocío University Hospital (Sevilla, Spain) and QuirónSalud Sagrado Corazón Hospital (Sevilla, Spain). Maternal age ranged from 18 to 47 years. Exclusion criteria for all pregnant women included the presence of known severe medical conditions, clinical infections, and the use of prescription medications or antibiotics within 2 months prior to study enrolment. This study was conducted in accordance with the principles of the Declaration of Helsinki. The study protocol was reviewed and approved by the research ethics committee (REC) (ID/Number: 0452-N-22). Written informed consent was obtained from all study participants.

AF samples were collected from women at two different stages of pregnancy: (1) A total of 127 AF samples were collected via amniocentesis between gestational weeks 13 and 33 from 123 pregnant women (including 4 diamniotic pregnancies). The amniocentesis cohort included 56 pregnancies with normal foetuses and 71 pregnancies with foetal anomalies; (2) A total of 27 AF samples were collected at term delivery (>37 weeks’) from 25 women scheduled for elective caesarean sections (including 2 diamniotic pregnancies) (**Fig. 1**).

Negative controls were implemented at each stage of the study to address potential contamination. These consisted of extraction process controls using sterile saline solution collected concurrently with amniocentesis or caesarean sections, control of inoculation tools and culture media by inoculating saline solution onto agar plates under the same conditions as the samples, incorporation of sample-free DNA extraction controls in all nucleic acid extractions, and template-free PCR amplification controls.

Inclusion criteria for sample analysis included the absence of blood contamination, and a minimum sample volume of 5 ml. As a result, 12 samples were excluded from the study.

**Table 4** provides a summary of maternal and foetal characteristics, including maternal age, gestational age at the time of sample collection, and pregnancy data.

**Table 4.**
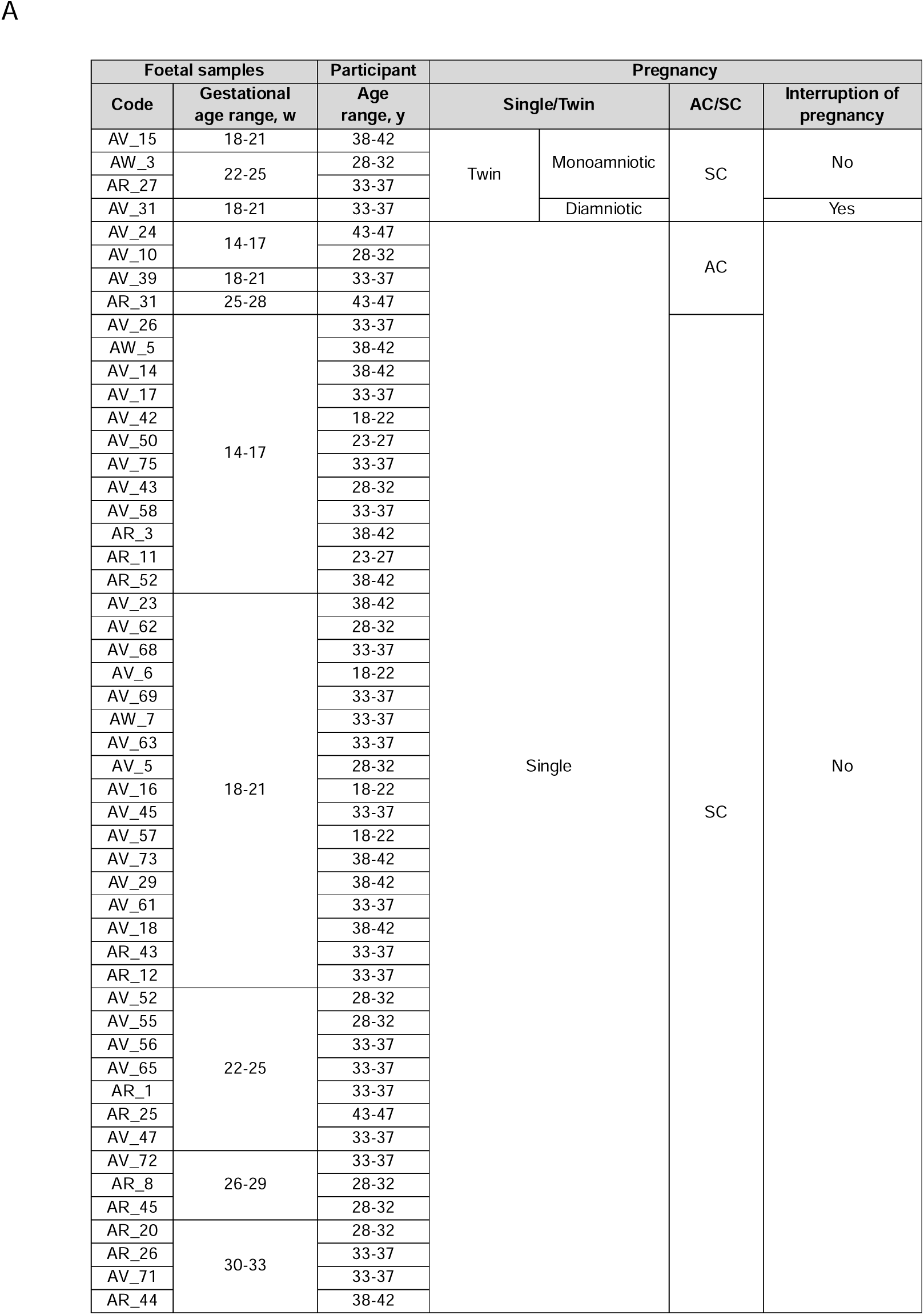
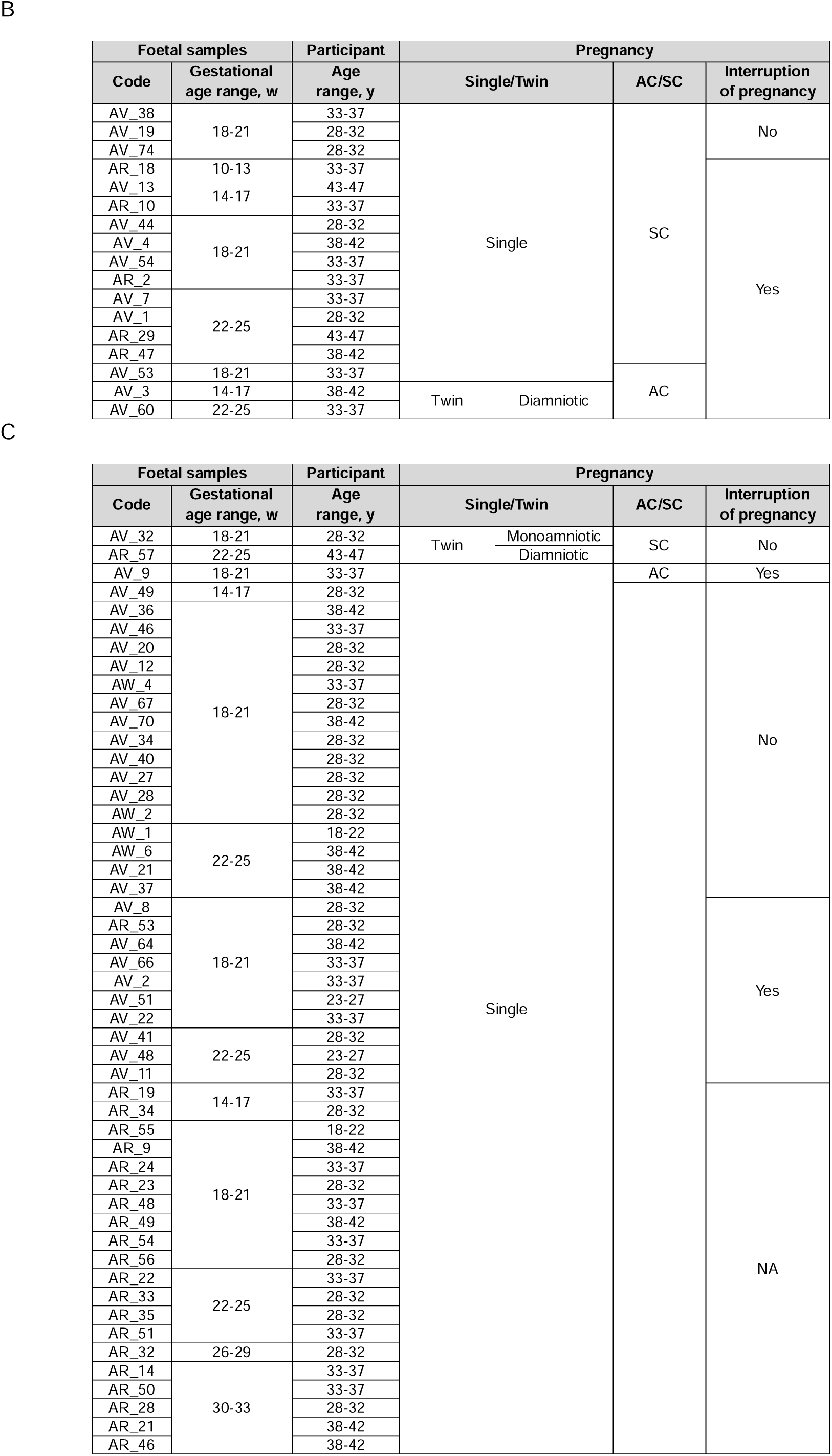
Clinical characteristics of pregnant women undergoing amniocentesis. **(A)** Pregnancies with normal foetuses. **(B)** Pregnancies with foetuses diagnosed with chromosomopathies. **(C)** Pregnancies with foetuses presenting malformations. Most samples corresponded to deliveries at a gestational age of over 37 weeks’, except for AV_46 (34 weeks’), AV_73 and AV_56 (35 weeks’), and AV_32 and AW_6 (36 weeks’). AC, assisted conception; NA, not available; SC, spontaneous conception; VD, vaginal delivery; w, week; y, year.

### Amniotic fluid sample collection

AF samples (n=154) were collected aseptically and stored atL−L80 °C until analysis. The collection of AF samples was conducted as follows: (1) during amniocentesis in the second trimester after abnormal findings on foetal ultrasound or biochemical marker analysis, by inserting a needle through the abdomen into the uterus to remove a 5-10 mL AF sample and, (2) during elective (planned, with no ongoing labor) caesarean delivery, at the time of caesarean section by aspirating through intact amniotic membranes using a sterile probe and 20-mL syringe.

To mitigate the risk of contamination, strict protocols were implemented throughout the collection and transport process. These protocols were based on aseptic techniques, the use of sterile single-use equipment, and immediate refrigeration of samples to minimise environmental exposure. Personnel followed standardised procedures, including wearing protective equipment and operating in controlled environments. This meticulous approach ensured the integrity of the AF samples, preserving their suitability for subsequent microbial analysis.

### Optimization of procedures for microbial culture analysis of AF samples

#### Concentration of AF samples

Each AF sample was divided into 2.5 mL aliquots. Aliquots were concentrated 2-fold, 3-fold, 4-fold, or 5-fold by centrifugation (Heraeus Biofuge Pico, Thermo Scientific). To optimise centrifugation parameters without compromising bacterial viability, AF concentrates were centrifuged at 1,250 ×Lg, 3,850 ×Lg, 9,500 ×Lg, and 13,250 ×Lg, for durations ranging from 0 to 15 minutes at room temperature. Supernatant was then discarded.

#### Culture media and incubation conditions

Pellets (100 µL) of each AF concentrate samples were inoculated and streaked in duplicate onto four media types: Brain Heart Infusion (BHI) Agar (Millipore, Merck, Darmstadt, Germany), Columbia Blood (CB) Agar (VWR Chemicals, Barcelona), Gifu Anaerobic Medium (GAM) Agar (Nissui Pharmaceutical, Japan), and MacConkey (MC) Agar (Millipore, Merck, Darmstadt, Germany). Culture media were selected based on previous studies to isolate microorganisms from AF samples (Collado et al., 2016; Romero et al., 2019; Theis et al., 2019; Winters et al., 2022). Plates were incubated at 37°C under aerobic conditions (BHI, CB, and MC) and anaerobic conditions (BHI, CB, GAM and MC), using AnaeroGen 2.5 L gas-generating sachets (Thermo Scientific, Oxoid), which create an anaerobic atmosphere (<1% O_2_, ∼10–15% CO_2_) inside an Oxoid jar. Plates were inspected daily for bacterial growth for up to two weeks under aerobic conditions and for four weeks under anaerobic conditions.

To maintain stringent anaerobic conditions throughout the incubation period, the anaerobic environment was continuously monitored, and gas-generating sachets were replaced on a weekly basis.

#### Pre-enrichment step

To recover low-abundance or fastidious organisms, a 200 µL aliquot of each 5-fold concentrate was inoculated into 4 mL sterile BHI broth and incubated aerobically at 37°C for 48 h with gentle agitation (200 rpm). After enrichment, 100 µL was sub-cultured onto BHI, CB, GAM, and MC plates as described above.

### Bacterial culture and 16S rRNA gene sequencing

For bacterial culture from AF samples, BHI, CB, GAM and MC Agar were used. Samples were plated and incubated at 37°C under either aerobic or anaerobic conditions in order to maximize the detection of cultivable microorganisms. Each strain was subcultured for an additional 48 hours under the same conditions, and DNA extracted using the GeneJET Genomic DNA Purification Kit (Thermo Scientific S.L., K0721). PCR was subsequently performed on each isolated bacterial colony using universal bacterial primers targeting the 16S ribosomal RNA (rRNA) gene (27F/1492R). Negative control samples included PCR amplification reagents and DNA-free water subjected to the extraction protocol. Gel electrophoresis confirmed the presence of amplicons of the expected size (∼1500bp). PCR products were purified and sequenced by STAB VIDA (Portugal) using pairwise Sanger sequencing. Sequence chromatograms were inspected manually using ChromasPro 2.2 to ensure accuracy of base calls, and the resultant sequence was entered into the National Center for Biotechnology Information (NCBI) Basic Local Alignment Search Tool (BLAST) to determine bacterial taxonomy. Sequences with more than 99% similarity and 99% query coverage were considered the same species.

### Assessment of the capacity of AF samples for bacterial growth

AF samples were individually inoculated with four bacterial species (*Bacillus subtilis*, *Cutibacterium acnes, Micrococcus luteus* and *Staphylococcus epidermidis*) in sterile microcentrifuge tubes and incubated at 37°C on a shaker at 200 rpm under both, aerobic and anaerobic conditions, to ensure uniform bacterial growth. Each microorganism was cultured in liquid BHI medium until reaching an optical density (OD) of 0.5, corresponding to the exponential growth phase, and a 1:10 dilution of the culture was used to inoculate the AF samples. To assess bacterial growth, absorbance at 600 nm was measured at time 0 and after different incubation times (6, 12, 24, 48, 72, 96 hours) using Nunc cell culture plates (polystyrene 96-well plates, Thermo Scientific). Negative controls containing only water and culture medium without sample were included and subjected to the same procedure as the inoculated samples. In order to quantify the colony-forming units (CFU) of the bacterial species that survive and proliferate in the AF after inoculation, these samples were cultured in BHI medium at 37°C under both aerobic and anaerobic conditions.

### Nucleic acid extraction and sequencing

DNA was extracted from each AF sample using two different protocols in parallel, with the aim of selecting the kit with the greatest yields and lowest number of contaminants: 1) QIAamp PowerFecal Pro (PFP) DNA kit (QIAGEN Pty Ltd, Hilden, Germany) and 2) QIAGEN MagAttract Microbial (MA) DNA kit (QIAGEN Pty Ltd, Hilden, Germany) on the Kingfisher DUO platform (Thermo Fisher Scientific) following the manufacturer’s instructions. A blank extraction control, consisting of DNA-free water, was included in each batch of extractions. The integrity and purity of the extracted nucleic acids were assessed via NanoDrop ND-1000 spectrophotometer and agarose gel electrophoresis, and DNA concentrations were quantified with a Qubit 4.0 Fluorometer (Thermo Fisher Scientific, Waltham, MA, USA).

DNA was analysed using two types of NGS techniques **(Fig. 6)**. First, shotgun metagenomic sequencing was employed using a NovaSeq 6000 system (Illumina, USA) through Novogene Europe (Cambridge, UK). Library preparation and sequencing generated 150 bp paired-end reads. Raw reads were processed to remove adapters, low-quality sequences, and ambiguous bases using the NovogeneAIT Genomics pipeline. The cleaned reads were subsequently used for bioinformatics analysis. Second, a 16S rRNA gene amplicon sequencing strategy was utilised, with amplicons obtained by PCR using asymmetric combinations of 5 forward primers and 24 reverse primers with barcodes (27F/1492R primer set), as previously described by Stinson et al. (2019a). PCR reactions were prepared using dsDNase-treated mastermixes (ArcticZymes) to minimise contaminant DNA. PCR cycling conditions included 35 cycles of amplification. Amplicons were visualised using a High Resolution Kit on a QIAXcel capillary gel electrophoresis system and pooled in equimolar concentrations based on band quantification within the target size range (1336-1743 bp). Subsequently, pooled samples were concentrated using NucleoMag NGS Clean-up and Size Select magnetic beads (Macherey Nagel) according to the manufacturer’s instructions and eluted in 75 μL volumes. 500 ng of purified DNA was used for library preparation at the Australian Genome Research Facility (AGRF). There, SMRTbell adapters were ligated to the barcoded PCR products, and the library was sequenced using the PacBio Revio system on a single SMRT cell.

**Figure 6.**
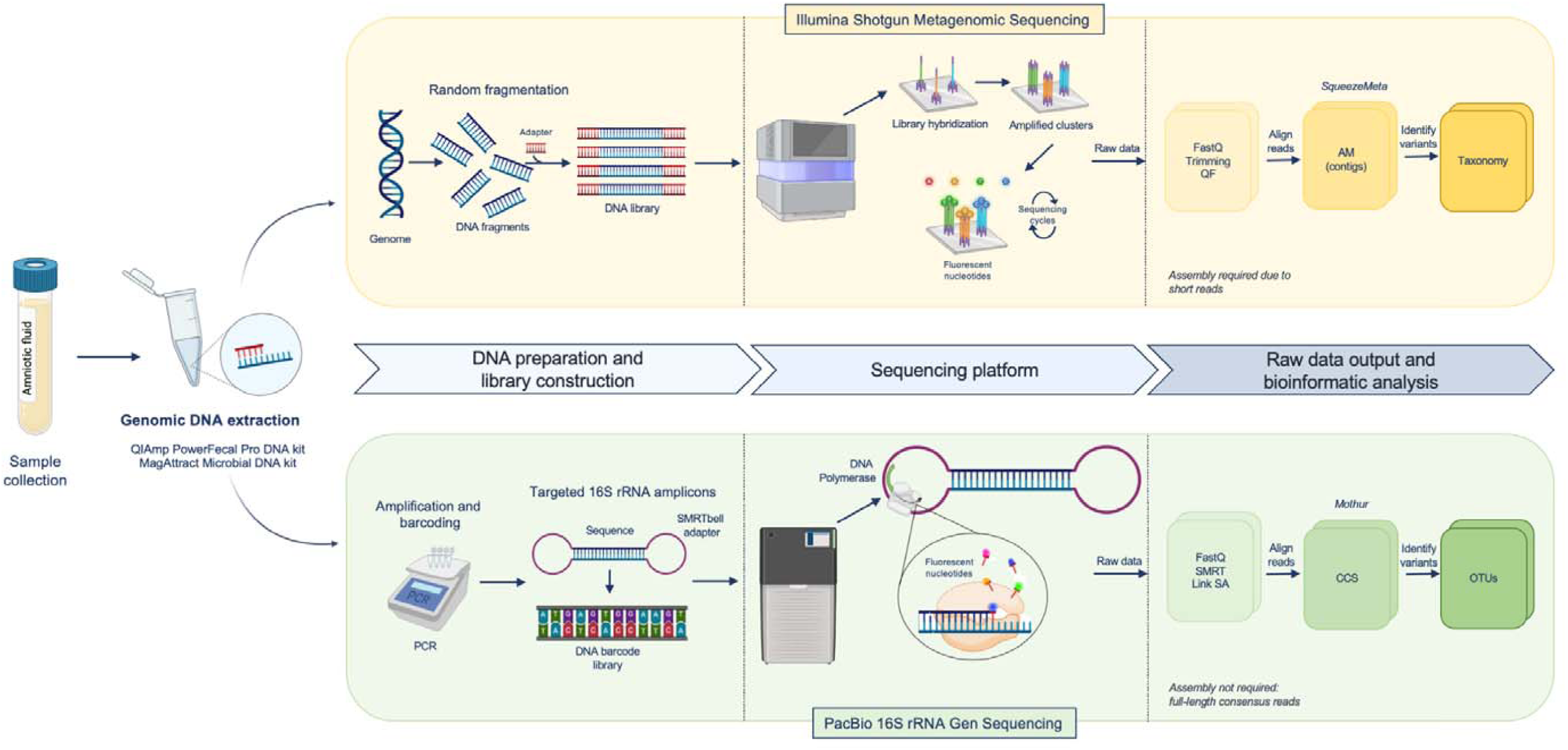
Workflow of NGS techniques for AF DNA samples with Illumina shotgun and PacBio 16S rRNA gene sequencing. AM, assembled metagenome; CCS, circular consensus sequencing; OTUs, operational taxonomic units; QF, quality filtering; SA, analysis; SMRT, single molecule real sequencing time.

### Detection of HBD-1, HBD-2, HBD-3, HNPs 1-3, and LL-37 by ELISA

A specific and sensitive enzyme-linked immunosorbent assay (ELISA) was used for the detection of AMPs, HBD-1, HBD-2 and HBD-3 in AF samples via the HBD-1, HBD-2 and HBD-3 kits from Aviscera Bioscience (Cat#SK00858–06, Santa Clara, CA). HNPs1-3 and LL-37 were detected using Human ELISA Kits from Hycult Biotech (Netherlands). All ELISA kits had been validated for use in prior studies (Espinoza et al., 2003, Varrey et al., 2018 and Para et al., 2020). AMP concentrations were calculated by interpolation from individual standard curves composed of purified defensins (0.97 to 125 pg/mL for HBD-1, 31.25 to 2,000 pg/mL for HBD-2, 39.5 to 2,500 pg/mL for HBD-3, 156 to 10,000 pg/mL for HNPs 1-3, and 0.1 to 156.25 ng/mL for LL-37). The sensitivity of the assay was 0.3 pg/mL for HBD-1, 7.0 pg/mL for HBD-2, 10.0 pg/mL for HBD-3, 19.46 pg/mL for HNPs 1-3 and 0.14 ng/mL for LL-37.

### Data analysis

#### Bacterial isolate 16S rRNA gene sequencing

16S rRNA gene sequences obtained from the isolated species were assembled and edited using ChromasPro 2.2. Phylogenetic and molecular evolutionary analyses were conducted using MEGA X (Kumar et al., 2018; Stecher et al., 2020). Multiple sequence alignment was performed with ClustalW (Thompson et al., 1994). Phylogenetic trees were constructed using the Neighbor-Joining (NJ) algorithm (Saitou and Nei, 1987). The bootstrap test was applied with 1000 replicates to estimate the reliability of the tree topology, and the percentage of replicate trees in which the associated taxa clustered together is shown next to the branches. Evolutionary distances were computed using the Kimura 2-parameter model (Kimura, 1980), expressed as the number of base substitutions per site. All ambiguous positions were removed for each sequence pair using the pairwise deletion option.

#### Shotgun metagenomic sequencing

Cleaned metagenomic reads generated by shotgun sequencing on a NovaSeq 6000 platform (Illumina, USA) were analysed using the SqueezeMeta pipeline (Tamames and Puente-Sánchez, 2019). Reads were assembled into contigs using the MEGAHIT assembler (Li et al., 2015), optimised for complex metagenomic datasets. Clean reads were then mapped back to the assembled contigs using Bowtie2 (Langmead and Salzberg, 2012) to estimate coverage and relative abundances. Contigs were grouped into genomic bins based on sequence composition and coverage profiles using a consensus approach combining MetaBAT (Kang et al., 2015) and COmbat CO-Normalization Using conTrols (COCONUT) (Sweeney et al., 2016). Taxonomic classification of contigs and bins was inferred from gene-level annotations. Functional profiling was derived from annotated genes to assess potential metabolic pathways and gene categories present in the metagenome. All downstream analyses and data visualization were performed using the SQMtools R package (Puente-Sánchez et al., 2020), enabling seamless exploration of the taxonomic and functional structure of microbial communities from SqueezeMeta outputs.

#### PacBio full-length 16S rRNA gene sequencing

PacBio raw sequence reads were initially filtered to retain only circular consensus sequences (CCS) with a minimum of three full passes and an accuracy threshold of 99.5%. Sequences were then demultiplexed according to the asymmetric barcoding strategy. Sequence data processing was performed using the Mothur software package (v.1.48.0) (Schloss et al., 2009). Reads were filtered by length, ranging from 1336 to 1743 bp, and further processed to exclude sequences containing homopolymers longer than nine bases. Alignment was performed against the SILVA reference alignment database (v138) (Glöckner et al., 2017). Chimeric sequences and those mapping to non-bacterial taxa were subsequently removed. Sequences were clustered into Operational Taxonomic Units (OTUs) by calculating pairwise distances, followed by clustering with a similarity cut-off of 0.03 using the cluster.split command. Alpha diversity was assessed using richness (number of different OTUs) and Shannon diversity. Beta diversity was evaluated using PERMANOVA with Bray-Curtis distances, implemented within the vegan package of R. For differential abundance analysis, a prevalence threshold of 10% with a minimum count of three reads was applied. Taxonomic assignment of the OTUs of interest was performed using BLAST (Altschul et al., 1990; 1997), with sequence identity scores of >99% and >99% considered reliable matches at the genus and species levels, respectively.

Data processing and statistical analyses were performed using Microsoft Excel, SPSS software version 19 (SPSS Inc., IBM Corporation, Armonk, NY, USA), and R Studio (v. 2024.09.1+394) (R Core Team R, 2020). Data normality was assessed with the Shapiro-Wilk test. Non-parametric tests were applied when necessary, and corrections for multiple comparisons were performed. An unpaired two-tailed Student’s t-test was used to assess statistical significance, with a p-value of <0.05 considered significant. The Mann-Whitney U test was employed to compare quantitative variables in independent groups, while Wilcoxon signed-rank tests were applied for comparisons within dependent groups.

## Data Availability

Datasets generated during the current study can be found in the Sequence Read Archives, under accession PRJNA1288569.

## Online supplemental material

**Fig. S1**. Phylogenetic trees of the four most frequently isolated bacterial species from AF samples, constructed using the Neighbor-Joining method based on 16S rRNA gene sequences. **(A)** *Bacillus subtilis;* **(B)** *Cutibacterium acnes;* **(C)** *Micrococcus luteus;* **(D)** *Staphylococcus epidermidis*. Labels represent individual isolates, color-coded by collection center. Branch values indicate bootstrap support (1,000 replicates). **Fig. S2**. Bacterial growth dynamics in AF samples. **(A)** Optical density (O.D._600 nm_) profiles of AF samples incubated without bacterial inoculum under aerobic and anaerobic conditions. **(B)** Growth curves of *Bacillus subtilis* (yellow colour), *Cutibacterium acnes* (red colour)*, Staphylococcus epidermidis* (blue colour), and *Micrococcus luteus* (green colour) in AF samples along with the growth profile of uninoculated AF (grey colour), showing bacterial proliferation over time under aerobic and anaerobic conditions. **Table S1.** Concentration of AMPs (ng/ml) in AF samples from second and third trimester. AMPs, antimicrobial peptides; HBD-1, human β-defensin 1; HBD-2, human β-defensin 2; HBD-3, human β-defensin 3; HNPs1–3, human neutrophil peptides 1–3; LOQ, limit of quantification; ND, not detected.

## Acknowledgements

The authors are grateful to the gynecologists C. López, E. De la Hoz, A. López, MA. Delmas, F. Blanco, IA. Castillo, P. Luque, A. Maraví, A. Ramírez, J. Sánchez, P. Trillo, R. Escorial, MD. Sánchez, A. Barranco, B. Buezas, JL. Arias, C. Domínguez, B. Tripiana and JC. Santos at QuirónSalud Hospital for their collaboration in collecting AF samples during caesarean sections. Our sincere thanks also to the midwives F. Martínez and E. Flores for their management of AF samples collected from Valme Hospital and Virgen del Rocío Hospital, respectively. They also thank Danika Hope for her technical support with DNA extraction and 16S rRNA gene PCR. Special thanks are extended to the generous pregnant volunteers who enrolled in this study.

This work was supported by grants from the Federación de Asociaciones de Celíacos de España (FACE) (SUBN/2019/005) and US-15332/I+D+I FEDER Andalucía 2014-2020. María González-Rovira received a fellowship from the University of Seville, Spain (PIF del VI Plan Propio de Investigación y Transferencia II.2A).

## Ethics approval and consent to participate

This study was conducted in accordance with the principles of the Declaration of Helsinki. The study protocol was reviewed and approved by the Sevilla Sur Provincial Research Ethics Committee (ID: 0452-N-22) for samples from Virgen de Valme University Hospital (Sevilla, Spain), Virgen del Rocío University Hospital (Sevilla, Spain) and QuirónSalud Sagrado Corazón Hospital (Sevilla, Spain). Written informed consent was obtained from all the participants. The participants consented for the use of their data and/or samples for health-related research purposes.

## Authors’ contributions

Study concept and design: EM and MLM; acquisition of data: MG-R, MLM and AM-P; data analysis and interpretation: MG-R, MLM, KM and MP; technical and essential material support: JAS-B, LG-D, CM-P and JS; manuscript drafting: MG-R and MLM; critical revision of the manuscript: EM, MP and CS. All authors read and approved of the final manuscript.

