## Supplemental Figures and Tables for "An integrated analysis of amniotic fluid reveals a dynamic prenatal microbial landscape and regulatory axis"

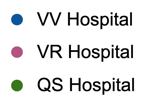
 A	B	C	D

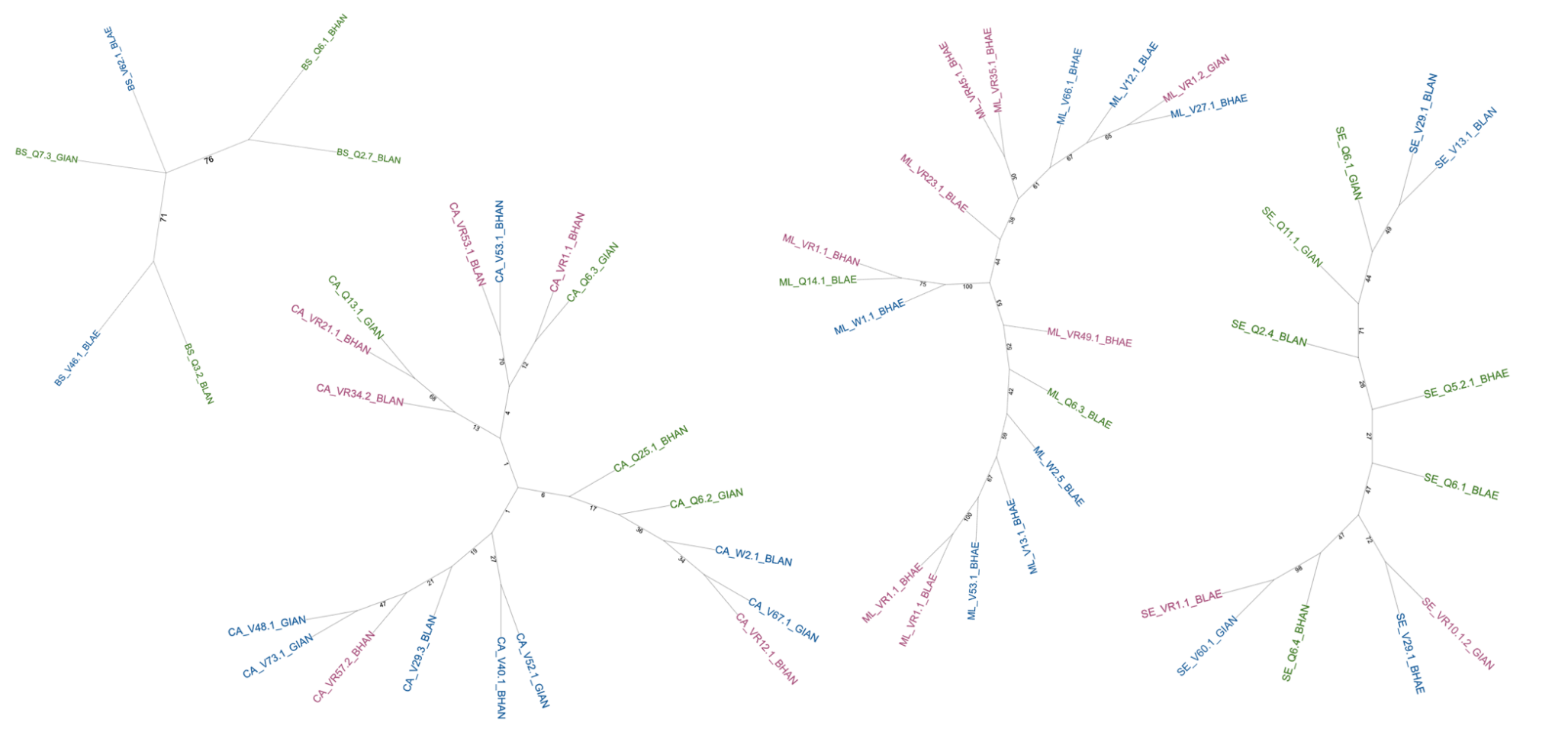

Figure S1. **Phylogenetic trees of the four most frequently isolated bacterial species from AF samples, constructed using the Neighbor-Joining method based on 16S rRNA gene sequences.** **(A)** *Bacillus subtilis;* **(B)** *Cutibacterium acnes;* **(C)** *Micrococcus luteus;* **(D)** *Staphylococcus epidermidis*. Labels represent individual isolates, color-coded by collection center. Branch values indicate bootstrap support (1,000 replicates).

A
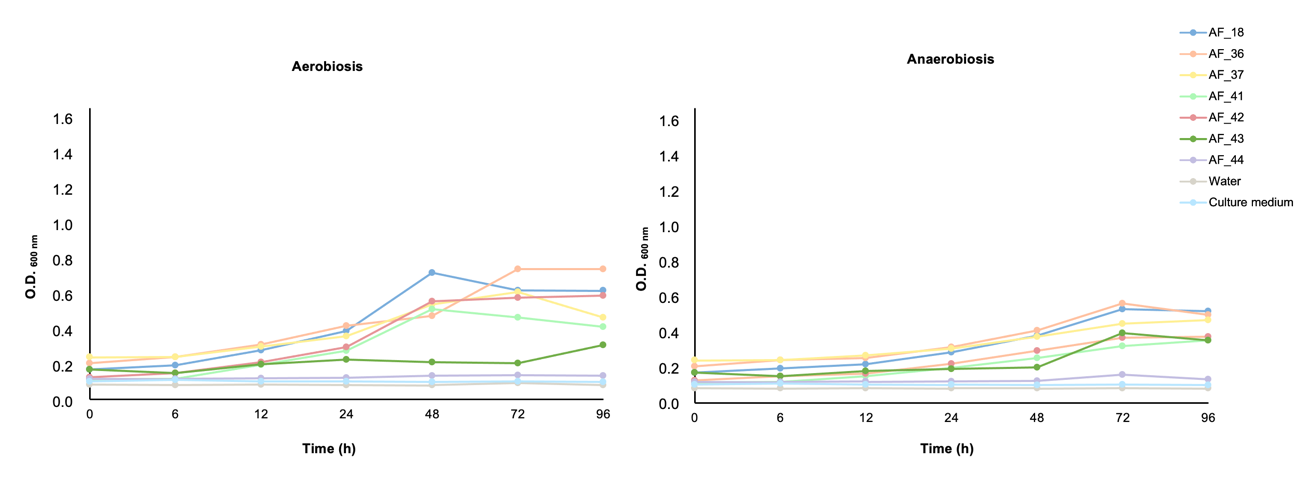

B

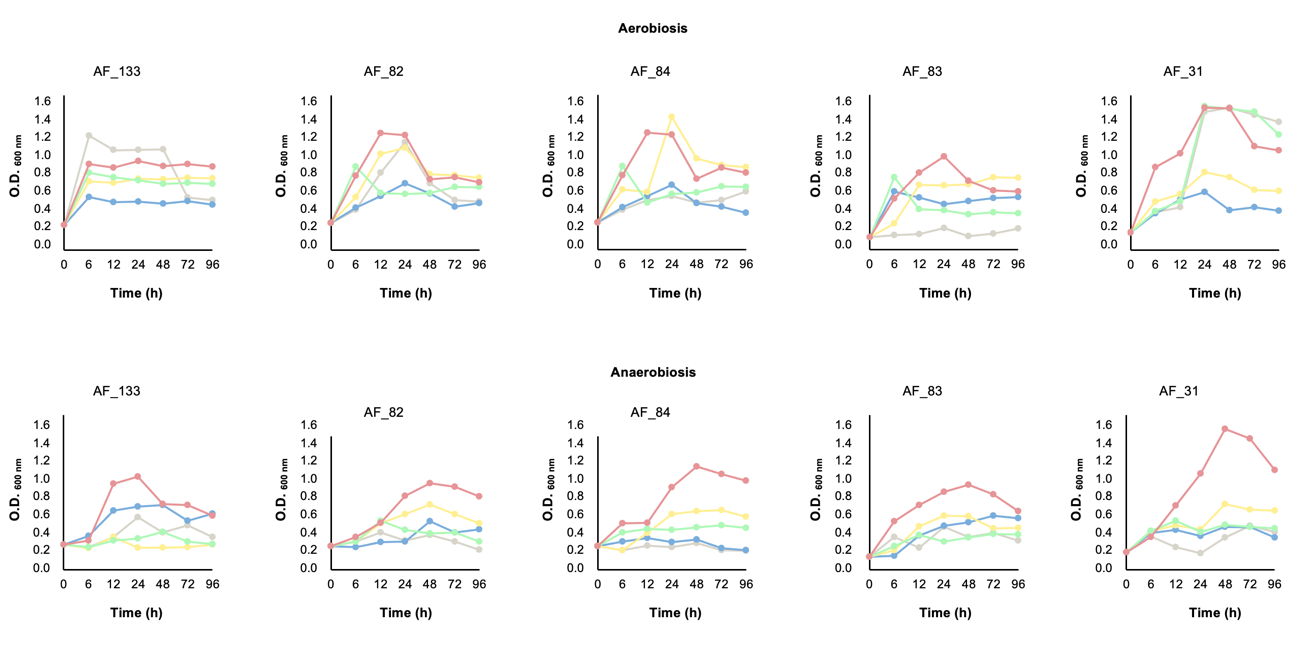

Figure S2. **Bacterial growth dynamics in AF samples.** **(A)** Optical density (O.D._600 nm_) profiles of AF samples incubated without bacterial inoculum under aerobic and anaerobic conditions. **(B)** Growth curves of *Bacillus subtilis* (yellow colour), *Cutibacterium acnes* (red colour)*, Staphylococcus epidermidis* (blue colour)*,* and *Micrococcus luteus* (green colour) in AF samples along with the growth profile of uninoculated AF (grey colour), showing bacterial proliferation over time under aerobic and anaerobic conditions.

Table S1. **Concentration of AMPs (ng/ml) in AF samples from second and third trimester**. AMPs, antimicrobial peptides; HBD-1, human β-defensin 1; HBD-2, human β-defensin 2; HBD-3, human β-defensin 3; HNPs1–3, human neutrophil peptides 1–3; LOQ, limit of quantification; ND, not detected.

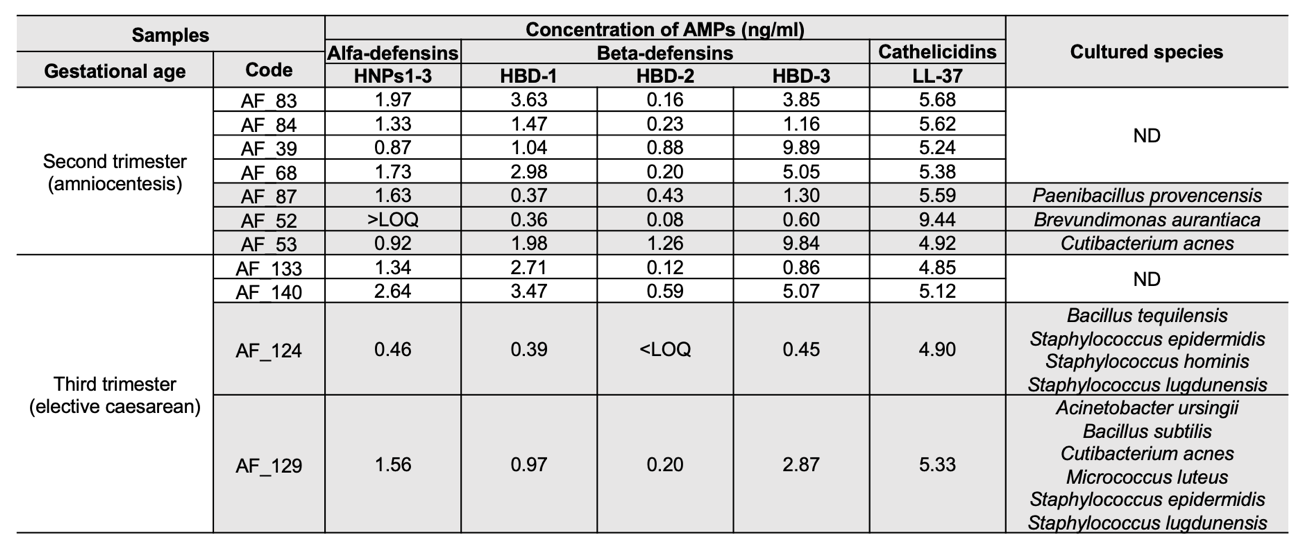
